# Causal relevance of new-onset type 2 diabetes mellitus and cancer risk in Chinese adults

**DOI:** 10.1101/2025.09.22.25336045

**Authors:** Mengying Wang, Owen Tipping, Bowen Liu, Derrick Bennett, Baiyong Shen, Siyi Zou, Shulin Zhao, Richard M. Martin, Matthew Sperrin, Andrew G. Renehan

**Affiliations:** Division of Cancer Sciences, School of Medical Sciences, Faculty of Biology, Medicine and Health, University of Manchester, Manchester, UK; Clinical Trial Service Unit and Epidemiological Studies Unit, Nuffield Department of Population Health, University of Oxford, Oxford, UK; Department of General Surgery, Pancreatic Disease Centre, Ruijin Hospital, Shanghai Jiao Tong University School of Medicine, Shanghai, China; Research Institute of Pancreatic Diseases, Shanghai Key Laboratory of Translational Research for Pancreatic Neoplasms, Shanghai Jiao Tong University School of Medicine, Shanghai, China; State Key Laboratory of Oncogenes and Related Genes, Institute of Translational Medicine, Shanghai Jiao Tong University, Shanghai, China; Department of Population Health Sciences, Bristol Medical School, University of Bristol, Bristol, UK; National Institute for Health Research (NIHR) Bristol Biomedical Research Centre, Bristol, UK; Centre for Health Informatics, Division of Informatics, Imaging and Data Sciences, School of Health Sciences, Faculty of Biology, Medicine and Health, University of Manchester, Manchester, UK; NIHR Manchester Biomedical Research Centre, Manchester, UK

**Author notes:** **Correspondence** Professor Andrew G. Renehan, Manchester Cancer Research Centre (MCRC), NIHR Manchester Biomedical Research Centre, Division of Cancer Sciences, School of Medical Sciences, Faculty of Biology, Medicine and Health, University of Manchester, Manchester, UK.

**Keywords:** Diabetes, Cancer, Longitudinal matching, China Kadoorie Biobank, Cohort study, Mendelian randomisation

## Abstract

**Background:** The prevalence of type 2 diabetes mellitus (T2DM) is increasing in China, and T2DM is linked to higher risk of several cancers, particularly obesity-related cancers (ORCs). Whether these associations are causal remains unclear due to potential biases.

**Methods:** We conducted a matched cohort study within the China Kadoorie Biobank (512,724 participants recruited 2004–08) to examine causal associations between new-onset T2DM and cancer incidence. To minimise bias, we: (i) included only incident T2DM cases, (ii) applied sex-specific sequential longitudinal matching, (iii) restricted analysis to pre-diagnosis body mass index (BMI), and (iv) accounted for detection time bias. Cox models stratified on the matched set estimated sex-specific hazard ratios (HRs) and 95% confidence intervals (CIs). Results were triangulated with two-sample Mendelian randomisation (MR).

**Findings:** After 1:3 matching, 8,657 men and 13,680 women with new-onset T2DM were retained, matched to 25,852 and 40,938 unexposed individuals, respectively. During follow-up to 31 Dec 2018, the incidence rate (IR) of total cancer was 1,365.1 per 100,000 person-years (95% CI 1,250.1–1,480.0) in men with T2DM versus 800.2 (749.7–850.7) in unexposed matches, and 864.7 (794.5–935.0) in women with T2DM versus 629.4 (594.7–664.1) in unexposed matches. T2DM was associated with increased risk of total cancer in men (HR 1.57, 95% CI 1.38–1.79) and women (HR 1.30, 95% CI 1.15–1.47). There was evidence of associations with liver (men HR 2.12, 95% CI 1.42–3.15; women HR 2.39, 95% CI 1.42–4.04) and pancreatic cancer (men HR 2.57, 95% CI 1.35–4.92; women HR 3.95, 95% CI 1.92–8.13). In MR, liability to T2DM was causally related with pancreatic cancer (pooled OR 1.08, 95% CI 1.02–1.15, P = 0.01), but not other cancers. Triangulation supported a causal link between T2DM and pancreatic cancer in East Asians.

**Interpretation:** These findings provide strong evidence for a causal relationship only between T2DM and pancreatic cancer risk in Chinese, while evidence for other cancer sites was weak or discordant.

**Funding:** China Scholarship Council; NIHR Manchester Biomedical Research Centre

**Research in context:** *Evidence before this study:* We systematically searched PubMed and Google Scholar for observational studies and/or Mendelian randomisation (MR) studies describing the associations between type 2 diabetes mellitus (T2DM) and incident cancer in Chinese. Existing Chinese cohort studies consistently suggested a modest excess risk in overall cancer incidence—driven mainly by liver, pancreas, colorectum, and in women, breast—but the strength of evidence was limited by recurring methodological shortcomings (e.g., inadequate covariate adjustment, immortal time and survivor biases, competing-risk bias). In MR studies among East Asians, except for pancreatic cancer, “counterintuitive” inverse associations were reported for several digestive cancers (e.g., stomach, liver, colorectum, oesophagus), raising concerns about potential flaws in the design of the source genome-wide association studies (GWAS). Overall, current evidence is constrained by substantial heterogeneity across studies, the likelihood of residual confounding (particularly from adiposity and lifestyle factors), other sources of bias, and possible publication bias. Robust, population-specific studies are needed to clarify the causal relationships between T2DM and site-specific cancer risks in China.

*Added value of this study:* To our knowledge, this is the first national-level prospective cohort study in China to employ a sex-specific, longitudinal matching strategy explicitly designed to minimise multiple sources of bias when evaluating the potential causal link between T2DM and cancer. Using rigorous methodology and triangulation with MR evidence, our findings provide causal support for an effect of T2DM on pancreatic cancer risk only. By contrast, many previously reported associations with other cancers, such as colorectum or female breast, are likely due to a range of biases.

*Implications of all the available evidence:* Our findings, together with prior research, indicate that the excess risks of most cancers observed in people living with T2DM are unlikely to be causal. The exception is pancreatic cancer, for which convergent evidence from both observational and genetic analyses supports a causal role of T2DM. In China—where diabetes prevalence is rising sharply— these results suggest that cancer prevention strategies should not treat T2DM as a broad carcinogenic exposure but rather focus on modifiable upstream determinants such as adiposity and lifestyle factors.

## Introduction

The prevalence of type 2 diabetes mellitus (T2DM) in China has surged dramatically in the past decades, increasing from approximately 22.56 million in 2000 to 140.87 million in 2021, with projections reaching 174.43 million by 2045.^1^ This is a substantial public health challenge, with a high economic burden associated with the management and treatment of T2DM in this country.^2^

A further concern is that there is evidence from many observational studies in European ancestry participants that T2DM is associated with higher cancer risks.^3^ However, it is unclear whether these associations are causal given multiple sources of bias. Common biases include (i) immortal time bias when prevalent T2DM cases are used; (ii) detection time bias where much of the increased risk is identified in the early time window after T2DM diagnosis through various co-detections of disease (for example, new incidental diagnosis of diabetes in the clinic during work-up for a cancer surgical resection); and (iii) confounding, for example, by key mutual risk factors like body fatness, commonly approximated by body mass index (BMI).^4^ Moreover, increased BMI is associated with at least 13 cancer types,^5^ but many observational studies in individuals with T2DM show an absence of an expected dose-response relationship for obesity-related cancers (ORCs).^6^ This is most likely due to weight loss associated with presentation and initial management of T2DM. Additionally, evidence from Mendelian randomisation (MR) studies do not support a causal relationship of the observed associations of T2DM on most cancer risks, except for pancreatic, cervical, and endometrial cancers.^6^ There are caveats that make it is difficult to generalise MR results across different populations with distinct genetic makeups or gene-environment interactions;^7^ and most well-powered genome-wide association studies (GWAS) of T2DM susceptibility were conducted in European ancestry populations.

In the Chinese population, observational studies have also identified T2DM as a risk factor for several cancer types including liver, pancreatic, colorectal, endometrial, and postmenopausal breast cancers.^8–11^ The study designs were either diabetes cohorts standardised against the Chinese population^8–10^ or cohort studies using prevalent T2DM at baseline^11^ – these are susceptible to immortal time bias. Cohort studies also encounter problems of adjusting for BMI measured after T2DM diagnosis. This may not reflect the pre-existing adiposity status of participants as weight changes frequently occur post T2DM diagnosis, influenced by metabolic alterations, intentional weight loss efforts, and treatments aimed at managing blood glucose levels.^12^ Thus, pre-T2DM BMI measurements might provide a more accurate representation of long-term adiposity exposure.

In this study, within the China Kadoorie Biobank (CKB), we aimed to assess the causal relevance of new-onset T2DM and cancer incidence. We designed a matched cohort study to reduce the above-mentioned biases by: (i) employing a sex-specific sequential longitudinal matching on age, study region, and most importantly, baseline BMI, to address confounding due to adiposity; (ii) including only new-onset T2DM cases, to minimise immortal time bias; (iii) restricting analysis in pre-diagnosis BMI, to mitigate reverse causation; and (iv) accounting for detection time bias. In addition, we utilised two-sample MR to strengthen causal interpretations.

This manuscript is one of two methodologically identical companion papers addressing this question in the Chinese population; the companion analysis uses UK Biobank data and is led by Owen Tipping and colleagues (preprint; DOI to be inserted).

## Methods

### Conventional observational analysis

#### Study population

The details of the study design of CKB have been reported elsewhere (http://www.ckbiobank.org/). The baseline survey recruited 512,724 participants aged 30–79 years between July 2004 and July 2008 from ten representative geographical regions in China. Detailed information on demographic characteristics and physical measures was collected by trained health staff at each site. Prior to commencement of CKB, ethics approval was obtained from the Oxford University Tropical Research Ethics Committee and the Chinese Centre for Disease Control and Prevention Ethical Review Committee, and all participants provided written informed consent. Further details on this cohort can be found in the *Cohort Profile* publication.^13^

#### Exposure

All information on non-fatal incident disease status was attained through linkage with established disease registries and the national health insurance system and coded using the 10^th^ Revision of the International Classification of Disease (ICD-10). The exposure of interest was new-onset diabetes (ICD-10: E11–E14), assuming any incident diabetes identified during follow-up was T2DM. We excluded participants with prevalent diabetes at baseline, determined through two methods as described in prior studies^14^ –– either physician-reported as such at baseline or screen-detected based on levels of random or fasting plasma glucose, detailed in supplementary figure 1.

#### Covariates

Sex assigned at birth was self-reported by participants and recorded at study entry. Anthropometric measurements were taken by trained health workers, with BMI calculated as weight (kg)/height (m^2^). The Working Group on Obesity in China suggests BMI thresholds of 24 kg/m^2^ for defining overweight and 28 kg/m^2^ for general obesity. Similarly, the Chinese Diabetes Society establishes waist circumference (WC) thresholds at 90 cm for men and 85 cm for women to define central obesity in the Chinese population.^15^ These criteria were adopted here to align with the characteristics of the study population.

We used key demographic and lifestyle factors recorded at baseline as covariates: physical activity (in metabolic equivalent task [MET] hours, continuous), WC (continuous), smoking and alcohol consumption (ever regular: YES/NO), education (≥6 years: YES/NO), and annual household income (≥35,000 yuan: YES/NO). For composite cancer risk estimates including liver cancer, we adjusted for cirrhosis/chronic hepatitis history (YES/NO). The selection of these covariates was informed by existing background knowledge of potential confounders in this context. At study entry, participants completed questionnaires under professional guidance, ensuring complete data collection.

#### Outcome measures

The primary outcome was first incident cancers (ICD-10: C00–C97). We examined 16 major cancer types reported in CKB, grouping them into composite outcomes: ORCs, digestive tract cancers, digestive accessory organ cancers, and ORCs in female reproductive organs (supplementary table 1). Postmenopausal breast cancer was defined as a diagnosis in women over 50 years due to the lack of updated menopause status.^16^ Since histological and anatomical details were unavailable, we could not differentiate between oesophageal adenocarcinoma (EAD) and squamous cell carcinoma (ESCC), or cardia gastric cancer (CGC) and non-cardia gastric cancer (NCGC). In China, > 85 % of oesophageal cancers are squamous-cell and two-thirds of gastric cancers are non-cardia,^17,18^ both of which have weak or inconsistent links to obesity; therefore cancers of the oesophagus (ICD-10: C15) and stomach (ICD-10: C16) were excluded from the ORC composite. The resulting ORC group captures the major adiposity-related sites that are prevalent in the Chinese population, rather than the full set of 13 sites listed by international agencies.^5^

Follow-up for all outcomes was administratively censored on 31 December 2018.

#### Matching

To reduce immortal time bias, we used a sequential longitudinal matching design.^19^ A 1:3 matching without replacement was implemented to enhance statistical power while ensuring most exposed individuals would be able to find at least one match.

Matching on baseline BMI was a key objective. The sequential longitudinal matching repeated the matching process throughout the follow-up period, starting from the date of first incident T2DM diagnosis and continuing enrolment whenever a new participant was clinically diagnosed with T2DM (i.e., the index date)(supplementary figure 2, A).^19^ All participants who remained on the study and had not yet been diagnosed with T2DM or any cancer at each index date were initially eligible for matching. The main effect of interest was analogous to the “per-protocol” effect, with matched individuals in the unexposed group being followed up until the diagnosis of T2DM or first primary cancer (whichever came first) for consistency with our study question (supplementary figure 2, B).

The overall eligibility of a potential unexposed match was defined by the following criteria: remained in study, remained undiagnosed with T2DM or cancer, from the same study region, at similar age on the index date with the exposed individual (± 3 years), and had similar baseline BMI with the exposed individual, defined by a caliper width of 0.25 times the standard deviation (SD).

#### Statistical analysis

In 1:3 matching, any T2DM case that found at least one suitable match was retained; therefore, the final analytic set included slightly fewer than three unexposed matches per case. To display covariate balance we applied pair-matching under the same algorithm and calculated the standardised mean difference (SMD) for each baseline variable; |SMD| < 0.10 denoted acceptable balance.^20^ Sex- and region-specific T2DM incidence rates were age-standardised to the baseline age distribution of the entire CKB population (age bands: < 40, 40–49, 50–59, 60–69, ≥ 70 years) to enable fair comparison across regions.

Cancer risks associated with new-onset T2DM were estimated using a Cox proportional hazards model, with time since diagnosis (or matched time for unexposed) as the time scale, stratified by the matched set. The model specifically estimated time-split coefficients for the exposure by incorporating an interaction term between exposure and follow-up time windows, which were divided into periods of within 12 months and beyond 12 months. This division was based on the distribution of Schoenfeld residuals plotted against follow-up time, to account for early detection time bias. Hazard ratios (HRs) and their 95% confidence intervals (CIs) for time windows exceeding 12 months were reported. A univariate model (Model 1) was initially fitted, complemented by a multivariable model adjusting for WC, smoking status, alcohol consumption habit, physical activity, highest education level, annual household income, and cirrhosis/chronic hepatitis history (Model 2). All analyses were conducted separately for men and women.

In competing risks modelling, non-cancer death and first cancers at sites other than the outcome of interest were treated as competing events. Fine–Gray subdistribution hazard models, stratified on the matched set and adjusted as in Model 2, produced subdistribution hazard ratios (sHRs) for the > 12 month window; these were compared with the cause-specific HRs from the main Cox analysis.^21^

Sensitivity analyses were performed as follows: (i) lower the BMI caliper to 0.10 × SD; (ii) restrict to women who never smoked and never consumed alcohol to minimise residual confounding; and (iii) repeat the analysis under a 1:5 matching ratio.

#### Mendelian randomisation

Two-sample MR utilising summary-level genetic data from publicly available sources were applied in the present study (supplementary figure 3). Specifically, genetic instruments for T2DM were extracted from the largest scale trans-ethnic meta-analysis until December 2024.^22^ From CKB, we derived genetic association data, adjusted for age, sex, and 10 genetic principal components for major site-specific cancers among 100,640 participants genotyped with the bespoke CKB Axiom array. The genotyped subset was assembled in two waves that deliberately over-sampled major incident cases of stroke, myocardial infarction and chronic obstructive pulmonary disease (COPD) but also included large random sets of participants, so its age-, sex- and region-specific distributions remain broadly representative of the parent cohort after weighting.^23^ Details in sources of genetic summaries for the traits used in the present study are presented in supplementary table 2. Single nucleotide polymorphisms (SNPs) that met the genome-wide statistical significance threshold (P < 5 × 10^−8^) and were retained after linkage disequilibrium (LD) clumping (R^2^ = 0.001, distance = 10 Mb) were proposed as initial genetic instruments for T2DM (N = 193). These selected SNPs altogether explained approximately 38.9% variance associated with T2DM. The details for calculations in R^2^, F-statistics, and statistical power are provided in supplementary text 1. Detailed information for SNPs is presented in supplementary table 3. The inverse-variance weighted (IVW) method was used as the main analytic approach because, when all instruments are valid (or pleiotropy is balanced), it is the most efficient and well-powered estimator of the causal effect. To probe robustness to horizontal pleiotropy, we prespecified a suite of sensitivity estimators with complementary assumptions: the weighted median (consistent if ≥ 50% of the weight comes from valid instruments), the weighted mode (robust when the largest cluster of instruments identifies the true effect), and MR-Egger (which can detect/adjust for directional pleiotropy under the InSIDE assumption, albeit with lower power). We additionally used MR-PRESSO to identify and correct for outlier variants indicative of horizontal pleiotropy. Further, to minimise the influence of pleiotropy via BMI, we applied multivariable MR to adjust for BMI. For interpretability, all ORs and 95% CIs were rescaled to represent the change in cancer risk per doubling in the odds of T2DM.^24^

All P values were two-sided, with P < 0.05 considered as statistically significant; data analyses were carried out using R, version 4.2.1(R Foundation) using the following packages: *tidyverse* (version 4.2.2), *survival* (version 3.4.0), *cmprsk* (version 2.2.12), *TwoSampleMR* (version 0.6.8), *MVMR* (version 0.4.1), *ieugwasr* (version 1.0.1), *metafor* (version 4.6.0) and *forestplot* (version 3.1.6).

## Results

### Overview of eligible participants for matching

Among participants who were diabetes-free at baseline (N=482,424), those with a history of cancer (N=2,284) or with missing values in BMI (N=2) were further excluded, and the remaining participants were initially eligible for matching (supplementary figure 4). Amongst the 197,656 men and 282,482 women eligible for matching, 9,206 (4.7%) men and 14,392 (5.1%) women were diagnosed with new-onset T2DM during the follow-up (regardless of cancer diagnoses), with a mean (SD) diagnostic age of 62.4 (10.2) years in men and 62.6 (9.6) years in women, respectively (supplementary table 4). The age-standardised incidence rate of T2DM showed large variations across the ten study regions in both sexes, e.g., the incidence rate in Zhejiang was more than three times higher than that in Gansu (supplementary figure 5).

### Associations of new-onset T2DM with baseline characteristics

Generally, individuals with new-onset T2DM were older, had lower education levels, higher family wealth, and higher rates of obesity at baseline. Regarding lifestyle factors, they were found to be less physically active and more prone to tobacco or alcohol use, albeit the differences were minor. Notably, the prevalence of a history of cirrhosis or chronic hepatitis was slightly higher in participants with new-onset T2DM (supplementary table 4).

### Construction of the analytic set

Following matching, the distribution of baseline BMI was well balanced. The remaining covariates reached a satisfactory level of balance (with |SMD| < 0.10), despite not being factors on which matching was performed (**table 1**). An analytic set was constructed following 1:3 matching, with 37 men and 36 women in the exposed group who failed to find any match excluded. The distributions of age at T2DM diagnosis and baseline BMI in all exposed participants before matching were presented in supplementary figure 6 and 7, respectively. Typically, these “fail” cases were outliers in terms of age and(or) BMI (supplementary table 5).

**Table 1.** Balance check in covariates following paired exact matching.

| Characteristic | Men |  |  | Women |  |  |
| --- | --- | --- | --- | --- | --- | --- |
|  | New-onset<br>T2DM | Matched<br>unexposed | SMD<sup>c</sup> | New-onset<br>T2DM | Matched<br>unexposed | SMD<sup>c</sup> |
|  | (N=8,675) | (N=8,675) |  | (N=13,689) | (N=13,689) |  |
| Follow-up time, median (IQR), months | 48.9(24.8–81.1) | 50.0(26.0–81.5) | .. | 53.2(27.7–86.7) | 53.0(27.6–85.4) | .. |
| Index age, mean (SD), years | 62.0(10.1) | 61.9(10.1) | .. | 62.3(9.6) | 62.2(9.5) | .. |
| Body mass index, mean (SD), kg/m<sup>2</sup> | 25.5(3.4) | 25.5(3.3) | 0.010 | 25.8(3.6) | 25.7(3.6) | 0.012 |
| **Study region, N (%)** |  |  |  |  |  |  |
| Qingdao | 622(7.2) | 622(7.2) | .. | 957(7.0) | 957(7.0) | .. |
| Harbin | 1,186(13.7) | 1,186(13.7) | .. | 1,650(12.1) | 1,650(12.1) | .. |
| Haikou | 473(5.5) | 473(5.5) | .. | 746(5.4) | 746(5.4) | .. |
| Suzhou | 1,040(12.0) | 1,040(12.0) | .. | 1,634(11.9) | 1,634(11.9) | .. |
| Liuzhou | 894(10.3) | 894(10.3) | .. | 1,127(8.2) | 1,127(8.2) | .. |
| Sichuan | 912(10.5) | 912(10.5) | .. | 1,828(13.4) | 1,828(13.4) | .. |
| Gansu | 376(4.3) | 376(4.3) | .. | 543(4.0) | 543(4.0) | .. |
| Henan | 501(5.8) | 501(5.8) | .. | 993(7.3) | 993(7.3) | .. |
| Zhejiang | 1,645(19.0) | 1,645(19.0) | .. | 2,634(19.2) | 2,634(19.2) | .. |
| Hunan | 1,026(11.8) | 1,026(11.8) | .. | 1,577(11.5) | 1,577(11.5) | .. |
| **Other baseline covariates** |  |  |  |  |  |  |
| Waist circumference, mean (SD), cm | 88.0(9.9) | 87.0(9.9) | 0.096 | 84.4(9.7) | 83.5(9.7) | 0.097 |
| ≥ 6 years of education, N (%) | 4,887(56.3) | 4,867(56.1) | 0.005 | 4,530(33.1) | 4,649(34.0) | -0.018 |
| Annual household income ≥ 35,000 yuan, N (%) | 2,272(26.2) | 2,294(26.4) | -0.006 | 2,591(18.9) | 2,601(19.0) | -0.002 |
| Ever regular smokers<sup>a</sup>, N (%) | 6,477(74.7) | 6,335(73.0) | 0.037 | 662(4.8) | 620(4.5) | 0.015 |
| Ever regular alcohol drinkers<sup>b</sup>, N (%) | 3,364(38.8) | 3,508(40.4) | -0.034 | 367(2.7) | 419(3.1) | -0.023 |
| Physical activity, mean (SD), MET h/d | 20.8(15.0) | 21.3(14.9) | -0.035 | 19.3(12.8) | 19.7(12.8) | -0.038 |
| History of cirrhosis/chronic hepatitis, N (%) | 172(2.0) | 135(1.6) | 0.032 | 131(1.0) | 115(0.8) | 0.012 |

Abbreviations: IQR, interquartile range; MET, metabolic equivalent task; SMD, standardised mean difference.
<sup>a</sup> Ever regular smokers included former and current regular smokers. <sup>b</sup> Ever regular alcohol drinkers included former regular and current weekly drinkers. <sup>c</sup> An absolute value less than 0.10 denoted acceptable balance.

### Observational associations between new-onset T2DM and cancer risks post matching

We noticed some evidence for early detection time bias, given the crude risk ratios (RRs) for total cancers in exposed individuals compared to the matched unexposed were higher in the first year but remained relatively stable afterwards in both men and women (**table 2**). In addition, when fitting a Cox model with constant coefficient for the exposure, the Schoenfeld residuals plotted against follow-up time showed a similar pattern (supplementary figure 8). Therefore, we put a cutoff point at 12 months, and the HRs corresponding to the time window after 12 months were presented.

**Table 2.** Unadjusted total cancer IRs in new-onset T2DM and in the matched unexposed, and crude RRs by annual time windows since index date following 1:3 exact matching.

|  |  |  | **Men** |  | **Women** |  |
| --- | --- | --- | --- | --- | --- | --- |
|  |  |  | **New-onset T2DM** | **Matched unexposed** | **New-onset T2DM** | **Matched unexposed** |
|  |  |  | (N=8,657) | (N=25,852) | (N=13,680) | (N=40,938) |
| Since index date | 0–1 year | No. of cases | 147 | 195 | 139 | 205 |
|  |  | Crude IR (95% CI)<sup>a</sup> | 275.3(230.8–320.8) | 125.7(108.1–143.4) | 195.0(162.3–227.0) | 95.0(81.9–108.0) |
|  |  | Crude RR (95% CI) | 2.19(1.77–2.71) | 1(ref) | 2.05(1.65–2.54) | 1(ref) |
|  | 1–2 years | No. of cases | 79 | 151 | 90 | 200 |
|  |  | Crude IR (95% CI)<sup>a</sup> | 161.9(126.2–197.6) | 106.3(89.3–123.2) | 140.8(111.7–170.0) | 100.0(86.1–113.9) |
|  |  | Crude RR (95% CI) | 1.52(1.16–2.00) | 1(ref) | 1.41(1.10–1.81) | 1(ref) |
|  | 2–3 years | No. of cases | 61 | 124 | 86 | 197 |
|  |  | Crude IR (95% CI)<sup>a</sup> | 99.5(74.5–124.5) | 64.9(53.5–76.3) | 90.4(71.3–109.5) | 68.2(58.7–77.8) |
|  |  | Crude RR (95% CI) | 1.54(1.13–2.08) | 1(ref) | 1.33(1.03–1.71) | 1(ref) |
|  | 3–4 years | No. of cases | 67 | 131 | 66 | 148 |
|  |  | Crude IR (95% CI)<sup>a</sup> | 141.4(107.5–175.2) | 89.5(74.2–104.9) | 90.4(68.6–112.2) | 66.1(55.4–76.8) |
|  |  | Crude RR (95% CI) | 1.58(1.18–2.12) | 1(ref) | 1.35(1.01–1.80) | 1(ref) |
|  | 4–5 years | No. of cases | 43 | 123 | 53 | 138 |
|  |  | Crude IR (95% CI)<sup>a</sup> | 94.1(66.0–122.2) | 85.5(70.4–100.6) | 71.2(52.1–90.4) | 61.5(51.3–71.7) |
|  |  | Crude RR (95% CI) | 1.10(0.78–1.56) | 1(ref) | 1.17(0.85–1.61) | 1(ref) |

Abbreviations: IR, incidence rate; RR, risk ratio.
<sup>a</sup> Per 1,000 person-years.

The HRs, corresponding to the average effect post 12 months, revealed that new-onset T2DM was associated with higher total cancer risk in men (HR 1.57, 95% CI 1.38–1.78). Specifically, men with new-onset T2DM exhibited elevated risks for ORCs (HR 1.73, 95% CI 1.37–2.19), digestive tract organ cancers (HR 1.33, 95% CI: 1.06–1.67), and for combined liver and pancreatic cancers (HR 2.23, 95% CI 1.59–3.11), as detailed in **table 3**. Women with new-onset T2DM had higher risks for total cancer (HR 1.29, 95% CI 1.14–1.46), ORCs (HR 1.34, 95% CI 1.10–1.62), and for combined liver and pancreatic cancers (HR 2.94, 95% CI 1.94– 4.45), aligning with the patterns observed in men. However, T2DM had no significant effect on the risk for ORCs in reproductive organs, including endometrial, ovarian, and postmenopausal breast cancers (HR 0.90, 95% CI 0.68–1.20) (**table 3**).

**Table 3.** Associations of new-onset T2DM with composite cancer risks following 1:3 exact matching.

|  |  | Men |  | Women |  |
| --- | --- | --- | --- | --- | --- |
|  |  | New-onset T2DM | Matched unexposed | New-onset T2DM | Matched unexposed |
| No. of participants |  | 8,657 | 25,852 | 13,680 | 40,938 |
| No. of person-years |  | 39,705 | 120,475 | 67,303 | 200,507 |
| *Total first primary cancers* |  |  |  |  |  |
| No. of cases |  | 542 | 964 | 582 | 1,262 |
| IR<sup>a</sup> |  | 1365.1(1250.1–1480.0) | 800.2(749.7–850.7) | 864.7(794.5–935.0) | 629.4(594.7–664.1) |
| HR (95% CI)<sup>b</sup> | Model 1<sup>c</sup> | 1.58(1.39–1.79) | 1(ref) | 1.32(1.17–1.49) | 1(ref) |
|  | Model 2<sup>d</sup> | 1.57(1.38–1.78) | 1(ref) | 1.29(1.14–1.46) | 1(ref) |
| *ORCs* |  |  |  |  |  |
| No. of cases |  | 176 | 279 | 222 | 520 |
| IR<sup>a</sup> |  | 443.3(377.8–508.8) | 231.6(204.4–258.8) | 329.9(286.5–373.2) | 259.3(237.1–281.6) |
| HR (95% CI)<sup>b</sup> | Model 1<sup>c</sup> | 1.81(1.44–2.28) | 1(ref) | 1.36(1.12–1.65) | 1(ref) |
|  | Model 2<sup>d</sup> | 1.73(1.37–2.19) | 1(ref) | 1.34(1.10–1.62) | 1(ref) |
| *Total cancers minus ORCs* |  |  |  |  |  |
| No. of cases |  | 366 | 685 | 360 | 742 |
| IR<sup>a</sup> |  | 921.8(827.4–1016.2) | 568.6(526.0–611.2) | 534.9(479.6–590.1) | 370.1(343.4–396.7) |
| HR (95% CI)<sup>b</sup> | Model 1<sup>c</sup> | 1.49(1.28–1.74) | 1(ref) | 1.30(1.12–1.50) | 1(ref) |
|  | Model 2<sup>d</sup> | 1.49(1.28–1.74) | 1(ref) | 1.27(1.09–1.47) | 1(ref) |
| *Digestive tract organ cancers* |  |  |  |  |  |
| No. of cases |  | 161 | 360 | 127 | 286 |
| IR<sup>a</sup> |  | 405.5(342.9–468.1) | 298.8(267.9–329.7) | 188.7(155.9–221.5) | 142.6(126.1–159.2) |
| HR (95% CI)<sup>b</sup> | Model 1<sup>c</sup> | 1.34(1.07–1.67) | 1(ref) | 1.18(0.93–1.51) | 1(ref) |
|  | Model 2<sup>d</sup> | 1.33(1.06–1.67) | 1(ref) | 1.14(0.89–1.46) | 1(ref) |
| *Digestive accessory organ cancers* |  |  |  |  |  |
| No. of cases |  | 104 | 117 | 75 | 100 |
| IR<sup>a</sup> |  | 261.9(211.6–312.3) | 97.1(79.5–114.7) | 111.4(86.2–136.7) | 49.9(40.1–59.6) |
| HR (95% CI)<sup>b</sup> | Model 1<sup>c</sup> | 2.36(1.71–3.25) | 1(ref) | 2.84(1.91–4.22) | 1(ref) |
|  | Model 2<sup>d</sup> | 2.23(1.59–3.11) | 1(ref) | 2.94(1.94–4.45) | 1(ref) |
| *ORCs in reproductive organs* |  |  |  |  |  |
| No. of cases |  | .. | .. | 79 | 262 |
| IR<sup>a</sup> |  | .. | .. | 117.4(91.5–143.3) | 130.7(114.8–146.5) |
| HR (95% CI)<sup>b</sup> | Model 1<sup>c</sup> | .. | .. | 0.92(0.69–1.21) | 1(ref) |
|  | Model 2<sup>d</sup> | .. | .. | 0.90(0.68–1.20) | 1(ref) |

Abbreviations: ORCs, obesity-related cancers; IR, incidence rate; HR, hazard ratio.
<sup>a</sup> Per 100,000 person-years. <sup>b</sup> The HR for follow-up time window over 12 months was presented. <sup>c</sup> Model 1 was stratified on the matched set. <sup>d</sup> Model 2 was stratified on the matched set and further adjusted for waist circumference, smoking status, alcohol consumption habit, physical activity, highest education level, annual household income, and cirrhosis/chronic hepatitis history (for composite cancer risks that encompassed liver cancer).

For site-specific cancer risks, in both men and women, significantly positive associations between new-onset T2DM and site-specific cancer risks were observed in liver (men HR 2.12, 95% CI 1.42–3.15; women HR 2.39, 95% CI 1.42–4.04) and pancreas (men HR 2.57, 95% CI 1.35–4.92; women HR 3.95, 95% CI 1.92–8.13). Notably, an elevated risk in lung cancer was observed in men (HR 1.55, 95% CI 1.17–2.05) but not in women (HR 1.15, 95% CI 0.87–1.52) (**table 4**). For the other cancer sites tested, we didn’t detect any significant signal.

**Table 4.** Associations of new-onset T2DM with site-specific cancer risks following 1:3 exact matching.

|  |  | **Men** |  | **Women** |  |
| --- | --- | --- | --- | --- | --- |
|  |  | **New-onset T2DM** | **Matched unexposed** | **New-onset T2DM** | **Matched unexposed** |
| No. of participants |  | 8,657 | 25,852 | 13,680 | 40,938 |
| No. of person-years |  | 39,705 | 120,475 | 67,303 | 200,507 |
| *Liver* |  |  |  |  |  |
| No. of cases |  | 68 | 86 | 43 | 68 |
| IR<sup>a</sup> |  | 171.3(130.6–212.0) | 71.4(56.3–86.5) | 63.9(44.8–83.0) | 33.9(25.9–42.0) |
| HR (95% CI)<sup>b</sup> | Model 1<sup>c</sup> | 2.29(1.57–3.34) | 1(ref) | 2.40(1.46–3.93) | 1(ref) |
|  | Model 2<sup>d</sup> | 2.12(1.42–3.15) | 1(ref) | 2.39(1.42–4.04) | 1(ref) |
| *Pancreas* |  |  |  |  |  |
| No. of cases |  | 36 | 32 | 33 | 33 |
| IR<sup>a</sup> |  | 90.7(61.1–120.3) | 26.6(17.4–35.8) | 49.0(32.3–65.8) | 16.5(10.8–22.1) |
| HR (95% CI)<sup>b</sup> | Model 1<sup>c</sup> | 2.54(1.38–4.68) | 1(ref) | 2.70(1.47–4.95) | 1(ref) |
|  | Model 2<sup>d</sup> | 2.57(1.35–4.92) | 1(ref) | 3.95(1.92–8.13) | 1(ref) |
| *Colorectum* |  |  |  |  |  |
| No. of cases |  | 62 | 143 | 64 | 145 |
| IR<sup>a</sup> |  | 156.2(117.3–195.0) | 118.7(99.2–138.2) | 95.1(71.8–118.4) | 72.3(60.5–84.1) |
| HR (95% CI)<sup>b</sup> | Model 1<sup>c</sup> | 1.39(0.98–1.98) | 1(ref) | 1.18(0.84–1.67) | 1(ref) |
|  | Model 2<sup>d</sup> | 1.33(0.93–1.91) | 1(ref) | 1.16(0.82–1.64) | 1(ref) |
| *Lung* |  |  |  |  |  |
| No. of cases |  | 116 | 202 | 101 | 212 |
| IR<sup>a</sup> |  | 292.2(239.0–345.3) | 167.7(144.5–190.8) | 150.1(120.8–179.3) | 105.7(91.5–120.0) |
| HR (95% CI)<sup>b</sup> | Model 1<sup>c</sup> | 1.52(1.15–2.00) | 1(ref) | 1.19(0.90–1.57) | 1(ref) |
|  | Model 2<sup>d</sup> | 1.55(1.17–2.05) | 1(ref) | 1.15(0.87–1.52) | 1(ref) |
| *Stomach* |  |  |  |  |  |
| No. of cases |  | 60 | 132 | 40 | 89 |
| IR<sup>a</sup> |  | 151.1(112.9–189.4) | 109.6(90.9–128.3) | 59.4(41.0–77.9) | 44.4(35.2–53.6) |
| HR (95% CI)<sup>b</sup> | Model 1<sup>c</sup> | 1.16(0.79–1.70) | 1(ref) | 1.16(0.75–1.78) | 1(ref) |
|  | Model 2<sup>d</sup> | 1.13(0.77–1.68) | 1(ref) | 1.11(0.72–1.71) | 1(ref) |
| *Breast (overall)* |  |  |  |  |  |
| No. of cases |  | .. | .. | 68 | 197 |
| IR<sup>a</sup> |  | .. | .. | 101.0(77.0–125.0) | 98.3(84.5–112.0) |
| HR (95% CI)<sup>b</sup> | Model 1<sup>c</sup> | .. | .. | 1.04(0.77–1.41) | 1(ref) |
|  | Model 2<sup>d</sup> | .. | .. | 1.01(0.73–1.38) | 1(ref) |
| *Postmenopausal breast* |  |  |  |  |  |
| No. of cases |  | .. | .. | 62 | 188 |
| IR<sup>a</sup> |  | .. | .. | 92.1(69.2–115.1) | 93.8(80.4–107.2) |
| HR (95% CI)<sup>b</sup> | Model 1<sup>c</sup> | .. | .. | 1.02(0.74–1.40) | 1(ref) |
|  | Model 2<sup>d</sup> | .. | .. | 1.02(0.74–1.41) | 1(ref) |

Abbreviations: IR, incidence rate; HR, hazard ratio.
<sup>a</sup> Per 100,000 person-years. <sup>b</sup> The HR for follow-up time window over 12 months was presented. <sup>c</sup> Model 1 was stratified on the matched set. <sup>d</sup> Model 2 was stratified on the matched set and further adjusted for waist circumference, smoking status, alcohol consumption habit, physical activity, highest education level, annual household income, and cirrhosis/chronic hepatitis history (for liver cancer).

### The effect of new-onset T2DM on absolute cancer incidence in the presence of competing risks

The cumulative incidence of total first primary cancers and competing non-cancer deaths before a potential cancer diagnosis (new-onset T2DM vs matched unexposed) was illustrated in supplementary figure 9. We also examined the censoring patterns in the two groups following 1:3 exact matching (supplementary table 6). While cumulative cancer incidence in the exposed group overrode that in the unexposed matches, competing risks from non-cancer deaths showed a similar trend, indicating that the HRs derived from a cause-specific hazard model could have overestimated the relative risk in cancer incidence in a real-world setting. In support of this, competing risks modelling resulted in attenuated sHRs compared to cause-specific hazard ratios from the main model by different degrees, but conclusions regarding effect directions remained unchanged (supplementary table 7).

### Sensitivity analyses

We applied a narrower caliper for BMI matching (0.1 SD, 1:3 matching), excluding 84 men and 74 women without suitable matches. Results remained consistent with the main analyses where a caliper equal to 0.25 SD was applied (supplementary table 8 and 9).

Using 1:5 matching, more exposed individuals remained unmatched, but results were generally consistent with 1:3 matching. An exception was a significantly positive association observed in lung cancer for women (HR 1.33, 95% CI 1.02–1.73) (supplementary table 10 and 11).

Among female participants eligible for matching at baseline (N=282,482), 61.2% were naïve to both cigarette and alcohol (N=172,903). In 9,083 exposed women with no history of tobacco or alcohol use, 31 lacked matches. Results aligned with the main analysis, showing significant associations between new-onset T2DM and total cancers (HR 1.27, 95% CI 1.10–1.46), ORCs (HR 1.39, 95% CI 1.10–1.76), and combined liver and pancreatic cancers (HR 2.05, 95% CI 1.34–3.12) (supplementary table 12). As shown in supplementary table 13, for site-specific cancers tested, a significantly positive signal was only observed for cancer in liver (HR 1.83, 95% CI 1.08–3.12) (the data for pancreatic cancer were not shown due to limited case number).

### Mendelian randomisation

Among the site-specific cancers tested, there was sufficient evidence of associations between genetic liability of T2DM and higher odds of pancreatic cancer (pooled OR 1.08, 95% CI 1.02– 1.15, P = 0.01). However, there was limited evidence for associations of genetically predisposed T2DM with cancers in colorectum, breast, endometrium, liver, lung, and stomach (**figure 1**). The results were consistent across sensitivity analysis (supplementary table 14) and after adjustment for BMI (supplementary figure 10). However, the estimation precision is low for some cancer types and the absence of significance might be due to the lack of statistical power (supplementary table 15).

**Figure 1.**
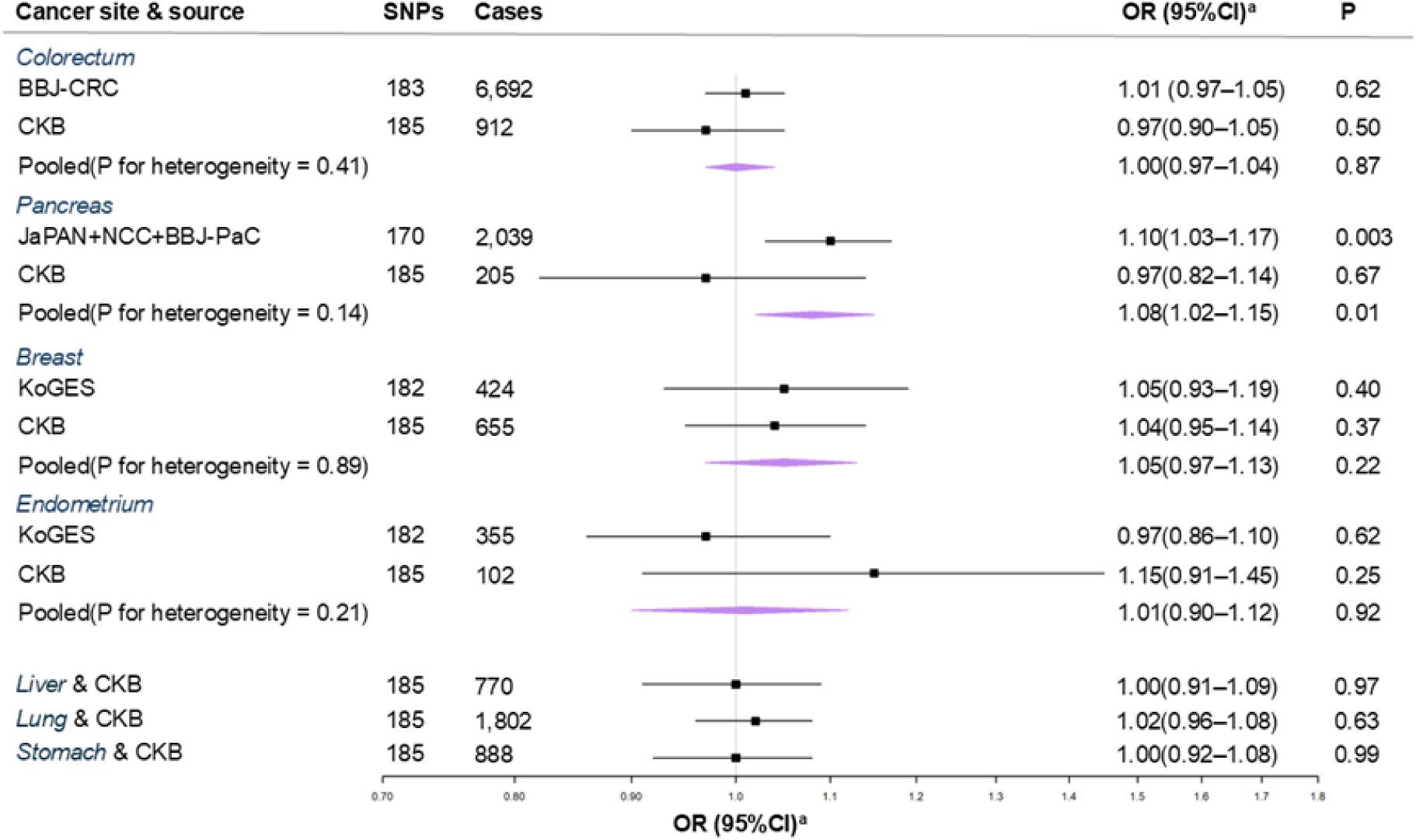
Univariable Mendelian randomisation for the associations of T2DM with major site-specific cancers in East Asians. Abbreviations: T2DM, type 2 diabetes mellitus; BBJ-CRC, Biobank Japan colorectal cancer cohort; CKB, China Kadoorie Biobank; JaPAN, Japan pancreatic cancer research; NCC, national cancer centre; BBJ-PaC, Biobank Japan pancreatic cancer cohort; KoGES, Korean genome and epidemiology study; OR, odds ratio; SNPs, single nucleotide polymorphisms. ^a^ Estimates indicating OR of cancer per doubling in odds of T2DM.

### Triangulation of evidence

For colorectal and pancreatic cancer, both methods showed directionally consistent effect sizes, with MR estimates—as expected—smaller in magnitude, leading to a concordant and precise interpretation. Breast and stomach cancers were also concordant, with both analyses indicating no association between T2DM and cancer risk; however, these estimates were imprecise due to limited statistical power. In lung cancer, a significant association was observed in men in the observational analysis, but not in women or in MR results. For liver cancer, the positive association seen in the observational analysis was not supported by MR findings (supplementary table 16).

## Discussion

### Summary of findings

Our analyses suggest that, after minimising key biases, new-onset T2DM is associated with higher risks of total cancer and, in particular, cancers of the liver and pancreas in Chinese adults. MR further supported a causal effect of genetic liability to T2DM on pancreatic cancer, whereas evidence for other sites was weak or inconsistent. Triangulating the observational and genetic results therefore supports a credible causal interpretation for pancreatic cancer but not for other cancers.

### Comparison with other studies and potential mechanisms

Earlier analyses in the CKB that relied on prevalent T2DM reported higher risks of total, liver, pancreatic, and breast cancer.^11^ In contrast, by prospectively identifying incident T2DM, matching chronologically to reduce immortal time bias, using pre-diagnosis BMI as a proxy for long-term adiposity, and accounting for early detection time bias via time-split coefficients, we again observed positive associations for liver and pancreatic cancers but no association for overall or postmenopausal breast cancer. The sex-specific patterning we observed—positive associations for lung and digestive-tract cancers in men but largely null estimates in women— likely reflects residual confounding by smoking and alcohol, exposures that are far more common among men in China.^13^ Results in women who were naïve to tobacco and alcohol were consistent with the main analyses, reinforcing this interpretation and motivating larger analyses among smoking- and alcohol-naïve men.

No association was observed for breast cancer (overall or postmenopausal), aligning with UK Biobank and Sister Study results.^26,27^ Limited cases precluded reliable estimates for endometrial and ovarian cancer, but the composite outcome (breast, endometrium, ovary) also showed null association. These hormone-related cancers appear driven more by adiposity than by downstream metabolic dysfunctions such as T2DM.^28^ Consistent with prior CKB findings^29^, our results suggest obesity, not T2DM per se, underlies elevated risk in these cancers.

For liver cancer, chronic hepatitis B/C virus (HBV/HCV) infection remains a dominant aetiology in China.^30^ We adjusted for baseline hepatitis/cirrhosis status, but subgroup analyses by viral infection were not feasible due to low prevalence data (HBV) and the absence of HCV information. The clinical implication is that identifying T2DM subgroups at particularly high liver-cancer risk—potentially defined by hepatitis status—merits attention in future work. For gastric cancer, our estimates could plausibly be biased upward by unmeasured Helicobacter pylori, which may be more prevalent in individuals with T2DM;^31^ limited site-specific case numbers further constrained precision. For colorectal cancer, our estimates—after accounting for early detection bias—do not indicate a robust association with new-onset T2DM, consistent with attenuation reported when early follow-up is excluded in prior study.^11^ Overall, the pattern emerging across digestive sites is that signals are strongest and most coherent for pancreas, weaker for liver (and potentially confounded by hepatitis), and largely null elsewhere after addressing early detection bias and confounding by adiposity and lifestyle.

Our observational findings are also broadly consistent with UK Biobank-derived results using similar methodology, showing strongest associations for liver and pancreatic cancer, and null effects for postmenopausal and endometrial cancers (Tipping et al. 2025, preprint, DOI to be inserted).

The MR results sharpen this picture. Across site-specific cancers with available summary statistics in East Asians, genetically proxied T2DM was associated with pancreatic cancer risk and showed little evidence for associations with colorectum, breast, endometrium, liver, lung, or stomach. The consistency between MR and our bias-minimised cohort analysis for pancreas supports a causal role of T2DM in pancreatic carcinogenesis. For other sites, discordance (e.g., observationally positive but MR-null for liver) suggests residual confounding in observational estimates or limited MR power, given modest case numbers for some cancers.

### Strengths and weaknesses

This study has several strengths that increase credibility. We focused exclusively on incident T2DM with covariates recorded at baseline, preserving correct temporality. Our sex-specific, sequential longitudinal matching on age, region, and baseline BMI directly targeted confounding by adiposity and improved exchangeability, while region matching helped mitigate geographic variation in unmeasured factors (dietary patterns, environmental exposures, and healthcare access). By explicitly modelling an early post-diagnosis window, we demonstrated the inflation of effect estimates within 12 months of T2DM onset—consistent with reverse causality and detection bias—and reported effects beyond that window. We also quantified subdistribution hazard ratios to reflect effect magnitudes in the presence of competing non-cancer deaths, which is more informative for clinical prediction and planning. Finally, triangulating with MR allowed us to separate likely causal from non-causal associations.

Limitations should be acknowledged. Power was limited for several sites due to the strict exposure definition and follow-up beginning at the index date, and some sites (e.g., thyroid, gallbladder) were absent or recorded without histological detail (e.g., oesophagus and stomach), preventing analyses by subtype. We lacked repeated measures of time-varying covariates prior to diabetes onset; reliance on a single baseline BMI may introduce measurement error and obscure adiposity trajectories that better capture lifetime exposure. Follow-up length constrained our ability to model risks across multiple post-diagnosis durations;^32^ prior evidence suggests duration of T2DM may modify liver and pancreatic cancer risks in Chinese populations, so extended observation in CKB will be informative.^33,34^ Residual confounding cannot be fully excluded—especially for smoking and alcohol in men—as implied by sex differences. Finally, MR power for some cancers was modest, so absence of genetic association does not rule out small effects.

### Implications

The broader implications are twofold. First, the convergence of observational and genetic evidence for pancreas argues that T2DM is part of the causal pathway for pancreatic cancer in East Asians. This strengthens the case for heightened vigilance and risk-adapted strategies among high-risk T2DM subgroups—particularly those with additional risk factors (e.g., older age at diabetes onset, obesity, heavy smoking or alcohol use, and chronic liver disease)— rather than blanket cancer screening for all people with T2DM. Second, for most other cancers, excess risks observed in people with T2DM likely reflect shared upstream determinants (notably adiposity and lifestyle) rather than diabetes per se, underscoring the primacy of prevention targeting weight, diet, and smoking/alcohol, alongside equitable access to routine, guideline-recommended screening in the general population.

### Unanswered questions and future research

Future research should refine estimates of cumulative pre-diabetes adiposity—e.g., incorporating BMI/WC trajectories from childhood and early adulthood—to reduce exposure misclassification and improve matched designs.^35,36^ Larger studies are needed to test effect modification by age at T2DM onset and BMI category, which were infeasible under caliper matching here but are clinically relevant for targeting interventions. Given the genetic and phenotypic heterogeneity of T2DM, integrating partitioned polygenic risk scores with phenotype-guided subtyping could reveal T2DM clusters with distinct cancer risks and mechanisms, improving risk stratification and informing mechanistically tailored prevention.^37,38^

## Conclusions

In conclusion, after addressing key sources of bias and triangulating with MR, we find robust evidence that T2DM increases pancreatic cancer risk in Chinese adults, with weaker and likely non-causal associations for most other sites. These results argue for targeted, risk-adapted cancer prevention within diabetes care and renewed emphasis on upstream determinants—particularly adiposity and smoking/alcohol—rather than treating T2DM as a broadly carcinogenic exposure.

## Data and resource availability

Bona fide researchers can apply for open-access CKB dataset by registering and applying at https://www.ckbiobank.org/data-access. Ancestry-specific GWAS summary for T2DM can be obtained from the DIAGRAM consortium (https://www.diagram-consortium.org/downloads.html). Summary-level data for colorectal cancer from BBJ-CRC sub-study and for BMI are publicly available on GWAS Catalog (study accession ID: GCST005591 and GCST90103751, respectively). Summary-level data for pancreatic cancer from the combined Japanese study are publicly available on the website of JaPAN consortium (http://www.aichi-med-u.ac.jp/JaPAN/current_initiatives-e.html). Summary-level data derived from CKB participants for the selected SNPs in the current study are available upon a reasonable request to the corresponding author.

## Contributors

MW: conceptualisation, methodology, project administration, formal analysis, writing—original draft. OT, BL, DB, BS, SZ, SZH, RMM and MS: conceptualisation, methodology, writing— review and editing. AGR: conceptualisation, project administration, funding acquisition, writing—review and editing. All authors reviewed and edited the final manuscript. All authors had full access to all the data in the study and had final responsibility for the decision to submit for publication.

## Acknowledgements

We thank all the participants, project staff, and the China National Centre for Disease Control and Prevention (CDC) and its regional offices for access to disease registries. MW gratefully acknowledges the financial support from the China Scholarship Council (CSC).

