## Supplementary text/figure/table for "Causal relevance of new-onset type 2 diabetes mellitus and cancer risk in Chinese adults"

Affiliations

**Supplementary Materials**

### Supplementary Text 1: Estimation of R^2^, F-statistics, and statistical power for summary-level MR

To evaluate the strength of genetic instruments for T2DM, we estimated the proportion of variance in the exposure explained by each SNP ($R^{2}$) and the corresponding *F-statistics*.

$R^{2}$ was calculated based on effect alle frequency ($\mathrm{EAF}$) and the SNP-exposure effect size ($\beta$) using the following equation:

$$R^{2}=2 \cdot EAF \cdot\left( 1-EAF \right)\cdot\beta^{2}$$

*F-statistics* were calculated for each SNP using the following equation:

$$F=\frac{{\left( N_{eff}-1 \right)\cdot R}^{2}}{1-R^{2}}$$

where $N_{eff}$ is the effective sample size. A value of *F* >10 was considered indicative of a strong instrument. Details in the calculated $R^{2}$ and *F-statistics* per SNP were presented in **supplementary table 3.**

To summarise instrument strength across all SNPs, a total *F-statistics* was calculated as:

$$F_{total}=\frac{\begin{aligned} \left( {N^{*}}_{eff}-K-1 \right)\cdot R_{total}^{2} \end{aligned}}{\begin{aligned} K\cdot(1-R_{total}^{2}) \end{aligned}}$$

where $K$ is the number of instruments, $R_{total}^{2}$ is the sum of individual $R^{2}$ values, and ${N^{*}}_{eff}$ is the harmonic mean of per-SNP effective sample sizes calculated as:

$$N_{eff}^{*}=\frac{n}{\sum_{i=1}^{n} \begin{aligned} \frac{1}{{\begin{aligned} (N_{eff}) \end{aligned}}_{i}} \end{aligned}}$$

$R_{total}^{2}$was then used in MR power calculations using an online calculator (https://sb452.shinyapps.io/power/).^1^

### Supplementary Table 1: Categorised cancer outcomes in the present study

| **Cancer site** | **ICD-10 codes** | **Digestive tract organs** | **Digestive accessory organs** | **ORCs** | | **ORCs in reproductive organs (women only)** |
| --- | --- | --- | --- | --- | --- | --- |
|  |  |  |  | **Men** | **Women** |  |
| All malignant neoplasms | C00–C97 |  |  |  |  |  |
| Lip, oral cavity and pharynx | C00–C14 | ● |  |  |  |  |
| Oesophagus | C15 | ● |  |  |  |  |
| Stomach | C16 | ● |  |  |  |  |
| Colon and rectum | C18–C20 | ● |  | ● | ● |  |
| Liver | C22 |  | ● | ● | ● |  |
| Pancreas | C25 |  | ● | ● | ● |  |
| Bronchus and lung | C33–C34 |  |  |  |  |  |
| Premenopausal breast | C50 |  |  |  |  |  |
| Postmenopausal breast | C50 |  |  |  | ● | ● |
| Cervix | C53 |  |  |  |  |  |
| Endometrium | C54.1 |  |  |  | ● | ● |
| Ovary | C56 |  |  |  | ● | ● |
| Prostate | C61 |  |  |  |  |  |
| Kidney | C64 |  |  | ● | ● |  |
| Bladder | C67 |  |  |  |  |  |
| Lymphoma | C81–C85 |  |  |  |  |  |
| Leukaemia | C91–C95 |  |  |  |  |  |

Abbreviations: M, men; W, women; ORCs, obesity-related cancers; ICD-10, the International Classification of Disease, 10^th^ Revision.

### Supplementary Table 2: Sources of genetic summaries for the traits used in the present study

| **Trait** | **Source** | **No. of participants** | **No. of cases** | **No. of controls** | **Reference** | **PMID** | **Sample source** |
| --- | --- | --- | --- | --- | --- | --- | --- |
| T2DM | DIAGRAM Consortium | 427,504 | 88,109 | 339,395 | Suzuki K, et al. (2024)^2^ | 38374256 | 40 GWAS in East Asians |
| BMI | TWB | 21,930 | NA | NA | Wong H, et al. (2022)^3^ | 35051171 | 21,930 Taiwanese from TWB |
| Colorectal cancer | BBJ-CRC study | 33,870 | 6,692 | 27,178 | Tanikawa C, et al. (2018)^4^ | 29471430 | BBJ-CRC Study (cases from BBJ, controls from three population-based cohorts) |
|  | CKB | 100,640 | 912 | 99,728 | NA | NA | 100,640 CKB participants with genotyping data at baseline |
| Pancreatic cancer | JaPAN+NCC+BBJ-PaC | 34,631 | 2,039 | 32,592 | Lin Y, et al. (2020)^5^ | 32581250 | Three Japanese studies: the JaPAN consortium, NCC, and the BBJ subcohort on pancreatic cancer (controls from four population-based cohorts) |
|  | CKB | 100,640 | 205 | 100,435 | NA | NA | 100,640 CKB participants with genotyping data at baseline |
| Breast cancer | KoGES | 46,330 | 424 | 45,906 | Nam K, et al (2022)^6^ | 36777999 | Three population-based prospective sub-cohorts in Korea |
|  | CKB | 57,625 | 655 | 56,970 | NA | NA | 57,652 female CKB participants with genotyping data at baseline |
| Endometrial cancer | KoGES | 46,330 | 355 | 45,975 | Nam K, et al (2022)^6^ | 36777999 | Three population-based prospective sub-cohorts in Korea |
|  | CKB | 57,625 | 102 | 57,523 | NA | NA | 57,652 female CKB participants with genotyping data at baseline |
| Liver cancer | CKB | 100,640 | 770 | 99,870 | NA | NA | 100,640 CKB participants with genotyping data at baseline |
| Lung cancer | CKB | 100,640 | 1,802 | 98,838 | NA | NA | 100,640 CKB participants with genotyping data at baseline |
| Stomach cancer | CKB | 100,640 | 888 | 99,752 | NA | NA | 100,640 CKB participants with genotyping data at baseline |

Abbreviations: NA, not applicable; T2DM, type 2 diabetes mellitus; GWAS, genome-wide association study; BMI, body mass index; TWB, Taiwan Biobank; BBJ-CRC, Biobank Japan colorectal cancer cohort; CKB, China Kadoorie Biobank; JaPAN, Japan pancreatic cancer research; NCC, national cancer centre; BBJ-PaC, Biobank Japan pancreatic cancer cohort; KoGES, Korean genome and epidemiology study.

### Supplementary Table 3: Genetic instruments for T2DM used in the present study

| **SNP** | **CHR** | **POS^a^** | **Effect allele** | **Other allele** | **BETA** | **SE** | **EAF** | **P** | **No. of cases** | **No. of controls** | **N_eff_** | **R^2^** | **F-statistic** |
| --- | --- | --- | --- | --- | --- | --- | --- | --- | --- | --- | --- | --- | --- |
| rs75372910 | 1 | 20688361 | T | C | -0.0451 | 0.0066 | 0.368 | 9.69E-12 | 88109 | 339395 | 247159 | 0.0009 | 234 |
| rs12119086 | 1 | 24371508 | A | G | -0.0677 | 0.0124 | 0.157 | 4.96E-08 | 42726 | 207363 | 112063 | 0.0012 | 300 |
| rs755249 | 1 | 39995074 | T | C | 0.0599 | 0.0088 | 0.1591 | 9.31E-12 | 88109 | 339395 | 247159 | 0.0010 | 238 |
| rs10749857 | 1 | 46248143 | T | C | -0.0602 | 0.0099 | 0.2955 | 1.00E-09 | 42529 | 203027 | 111309 | 0.0015 | 374 |
| rs12354253 | 1 | 51148711 | A | G | -0.0977 | 0.0112 | 0.0926 | 3.12E-18 | 87912 | 335059 | 246405 | 0.0016 | 397 |
| rs2269242 | 1 | 64108682 | A | G | -0.0583 | 0.0082 | 0.1901 | 9.83E-13 | 87912 | 335059 | 246405 | 0.0010 | 259 |
| rs11588753 | 1 | 146714427 | C | G | 0.051 | 0.0078 | 0.2163 | 5.18E-11 | 88109 | 339395 | 247159 | 0.0009 | 218 |
| rs670323 | 1 | 177868990 | A | G | 0.0577 | 0.0077 | 0.217 | 8.79E-14 | 88109 | 339395 | 247159 | 0.0011 | 280 |
| rs10797937 | 1 | 184017131 | A | G | -0.0404 | 0.0064 | 0.5403 | 2.43E-10 | 88109 | 339395 | 247159 | 0.0008 | 201 |
| rs6689629 | 1 | 204539291 | A | G | 0.0399 | 0.0066 | 0.6064 | 1.48E-09 | 87912 | 335059 | 246405 | 0.0008 | 188 |
| rs340874 | 1 | 214159256 | T | C | -0.0485 | 0.0065 | 0.616 | 8.84E-14 | 88109 | 339395 | 247159 | 0.0011 | 275 |
| rs348330 | 1 | 229672955 | A | G | -0.0555 | 0.0073 | 0.3119 | 4.15E-14 | 87912 | 335059 | 246405 | 0.0013 | 327 |
| rs7564711 | 2 | 639061 | T | G | -0.1022 | 0.011 | 0.0941 | 1.41E-20 | 88109 | 339395 | 247159 | 0.0018 | 441 |
| rs28680411 | 2 | 16237427 | A | G | 0.0425 | 0.0072 | 0.3984 | 3.88E-09 | 87950 | 337771 | 246579 | 0.0009 | 214 |
| rs1260326 | 2 | 27730940 | T | C | -0.0708 | 0.0064 | 0.5415 | 9.03E-29 | 88109 | 339395 | 247159 | 0.0025 | 617 |
| rs12712928 | 2 | 45192080 | C | G | 0.0545 | 0.0066 | 0.4017 | 1.83E-16 | 88109 | 339395 | 247159 | 0.0014 | 353 |
| rs243018 | 2 | 60586707 | C | G | -0.0578 | 0.007 | 0.3338 | 9.60E-17 | 88109 | 339395 | 247159 | 0.0015 | 368 |
| rs10184881 | 2 | 65684152 | A | G | 0.0515 | 0.0091 | 0.8493 | 1.39E-08 | 88109 | 339395 | 247159 | 0.0007 | 168 |
| rs3731600 | 2 | 120231070 | C | G | 0.1282 | 0.0191 | 0.9568 | 2.12E-11 | 83923 | 315971 | 234546 | 0.0014 | 336 |
| rs2167500 | 2 | 121370591 | A | C | 0.0374 | 0.0067 | 0.4901 | 1.93E-08 | 88109 | 339395 | 247159 | 0.0007 | 173 |
| rs4499362 | 2 | 149568396 | T | C | -0.043 | 0.0067 | 0.3611 | 1.85E-10 | 88109 | 339395 | 247159 | 0.0009 | 211 |
| rs75536691 | 2 | 165381518 | A | G | 0.1875 | 0.0214 | 0.9753 | 2.08E-18 | 85983 | 337970 | 244518 | 0.0017 | 419 |
| rs75179644 | 2 | 213687103 | T | C | 0.0746 | 0.011 | 0.8673 | 1.13E-11 | 88109 | 339395 | 247159 | 0.0013 | 317 |
| rs117809958 | 2 | 234191103 | A | T | 0.2703 | 0.0251 | 0.023 | 4.02E-27 | 87950 | 337771 | 246579 | 0.0033 | 814 |
| rs2067819 | 3 | 12359049 | A | G | -0.1069 | 0.0166 | 0.045 | 1.17E-10 | 85087 | 312200 | 236279 | 0.0010 | 243 |
| rs3860593 | 3 | 23102368 | A | G | -0.0587 | 0.0085 | 0.826 | 3.84E-12 | 88109 | 339395 | 247159 | 0.0010 | 245 |
| rs56979234 | 3 | 23436567 | A | G | -0.1115 | 0.0084 | 0.1801 | 4.09E-40 | 88109 | 339395 | 247159 | 0.0037 | 911 |
| rs704361 | 3 | 63883671 | T | G | -0.0829 | 0.0067 | 0.3514 | 1.48E-35 | 88109 | 339395 | 247159 | 0.0031 | 777 |
| rs12490891 | 3 | 84783138 | A | G | 0.0366 | 0.0065 | 0.3892 | 2.26E-08 | 88109 | 339395 | 247159 | 0.0006 | 158 |
| rs9860393 | 3 | 114966421 | A | G | 0.0446 | 0.0069 | 0.6891 | 8.07E-11 | 88109 | 339395 | 247159 | 0.0009 | 211 |
| rs6767514 | 3 | 123151762 | A | C | -0.053 | 0.0068 | 0.3474 | 7.09E-15 | 88109 | 339395 | 247159 | 0.0013 | 315 |
| rs13073553 | 3 | 125091755 | A | G | -0.0358 | 0.0065 | 0.5041 | 4.24E-08 | 87912 | 335059 | 246405 | 0.0006 | 158 |
| rs9873341 | 3 | 152387723 | T | C | 0.0474 | 0.0071 | 0.2782 | 2.05E-11 | 88109 | 339395 | 247159 | 0.0009 | 223 |
| rs9875793 | 3 | 170686573 | A | G | 0.061 | 0.0082 | 0.8151 | 1.31E-13 | 88109 | 339395 | 247159 | 0.0011 | 278 |
| rs4389513 | 3 | 185506857 | T | G | -0.129 | 0.0068 | 0.6882 | 2.95E-79 | 88109 | 339395 | 247159 | 0.0071 | 1778 |
| rs6444187 | 3 | 186674138 | C | G | -0.0421 | 0.0065 | 0.5252 | 6.93E-11 | 88109 | 339395 | 247159 | 0.0009 | 219 |
| rs34307738 | 3 | 195818265 | A | G | -0.0442 | 0.0066 | 0.3849 | 2.88E-11 | 88109 | 339395 | 247159 | 0.0009 | 229 |
| rs730831 | 4 | 1240299 | T | G | 0.099 | 0.0069 | 0.6806 | 3.93E-46 | 87912 | 335059 | 246405 | 0.0043 | 1058 |
| rs4865473 | 4 | 1782586 | A | G | -0.0694 | 0.0077 | 0.7062 | 1.51E-19 | 84327 | 328752 | 238303 | 0.0020 | 495 |
| rs4234731 | 4 | 6299914 | A | G | -0.1106 | 0.018 | 0.0613 | 7.77E-10 | 85754 | 336542 | 242392 | 0.0014 | 348 |
| rs10938398 | 4 | 45186139 | A | G | 0.0399 | 0.007 | 0.294 | 1.13E-08 | 88109 | 339395 | 247159 | 0.0007 | 163 |
| rs12499651 | 4 | 71854281 | A | G | 0.0713 | 0.0079 | 0.2065 | 2.26E-19 | 88109 | 339395 | 247159 | 0.0017 | 412 |
| rs117624659 | 4 | 85339618 | T | C | 0.2015 | 0.0209 | 0.9699 | 5.16E-22 | 87950 | 337771 | 246579 | 0.0024 | 587 |
| rs6813195 | 4 | 153520475 | T | C | -0.0773 | 0.0064 | 0.4907 | 1.35E-33 | 88109 | 339395 | 247159 | 0.0030 | 740 |
| rs6885132 | 5 | 14768092 | C | G | 0.0409 | 0.0065 | 0.555 | 2.86E-10 | 88109 | 339395 | 247159 | 0.0008 | 204 |
| rs878608 | 5 | 36246700 | T | C | -0.0563 | 0.0086 | 0.8282 | 6.62E-11 | 88109 | 339395 | 247159 | 0.0009 | 223 |
| rs72756138 | 5 | 50085981 | T | C | 0.056 | 0.0095 | 0.1276 | 4.15E-09 | 88109 | 339395 | 247159 | 0.0007 | 173 |
| rs3762991 | 5 | 52085403 | T | C | 0.0437 | 0.0069 | 0.3094 | 1.78E-10 | 88109 | 339395 | 247159 | 0.0008 | 202 |
| rs256904 | 5 | 55810305 | A | T | -0.0706 | 0.0064 | 0.5075 | 1.11E-28 | 88109 | 339395 | 247159 | 0.0025 | 617 |
| rs6453133 | 5 | 74692776 | A | G | 0.0413 | 0.0064 | 0.4881 | 9.69E-11 | 88109 | 339395 | 247159 | 0.0009 | 211 |
| rs6453436 | 5 | 78626464 | A | G | -0.0358 | 0.0064 | 0.4248 | 2.65E-08 | 88109 | 339395 | 247159 | 0.0006 | 155 |
| rs7721099 | 5 | 87936379 | T | C | -0.0385 | 0.0064 | 0.5378 | 2.10E-09 | 88109 | 339395 | 247159 | 0.0007 | 182 |
| rs3853212 | 5 | 95849348 | T | C | -0.0406 | 0.0064 | 0.5811 | 2.94E-10 | 88109 | 339395 | 247159 | 0.0008 | 199 |
| rs154120 | 5 | 111375061 | A | G | 0.0352 | 0.0064 | 0.5536 | 4.05E-08 | 88109 | 339395 | 247159 | 0.0006 | 151 |
| rs12657328 | 5 | 122651711 | T | C | 0.0632 | 0.0116 | 0.9073 | 4.73E-08 | 88109 | 339395 | 247159 | 0.0007 | 166 |
| rs451643 | 5 | 176511432 | T | G | 0.0531 | 0.0065 | 0.434 | 4.15E-16 | 85780 | 337056 | 242491 | 0.0014 | 343 |
| rs9379084 | 6 | 7231843 | A | G | -0.076 | 0.0084 | 0.2221 | 1.57E-19 | 88109 | 339395 | 247159 | 0.0020 | 494 |
| rs9350271 | 6 | 20683164 | A | G | 0.1947 | 0.0064 | 0.4218 | 1.06E-202 | 88109 | 339395 | 247159 | 0.0185 | 4656 |
| rs879882 | 6 | 31139452 | T | C | -0.0605 | 0.0064 | 0.5666 | 2.91E-21 | 88109 | 339395 | 247159 | 0.0018 | 445 |
| rs9265745 | 6 | 31302718 | A | G | 0.043 | 0.0064 | 0.4845 | 1.86E-11 | 87912 | 335059 | 246405 | 0.0009 | 228 |
| rs35800511 | 6 | 32636433 | A | G | 0.0485 | 0.008 | 0.7694 | 1.26E-09 | 83710 | 327595 | 236922 | 0.0008 | 206 |
| rs118012224 | 6 | 34205822 | T | C | 0.1156 | 0.0106 | 0.8927 | 9.75E-28 | 88109 | 339395 | 247159 | 0.0026 | 634 |
| rs742762 | 6 | 39046644 | A | C | 0.0801 | 0.0072 | 0.7132 | 1.15E-28 | 88109 | 339395 | 247159 | 0.0026 | 650 |
| rs62405419 | 6 | 50787459 | T | G | 0.0515 | 0.0072 | 0.2695 | 8.41E-13 | 88109 | 339395 | 247159 | 0.0010 | 258 |
| rs80196932 | 6 | 117996631 | T | C | 0.056 | 0.0079 | 0.7798 | 1.49E-12 | 88109 | 339395 | 247159 | 0.0011 | 266 |
| rs4273712 | 6 | 126964510 | A | G | -0.0568 | 0.0064 | 0.5319 | 5.64E-19 | 87117 | 337721 | 244667 | 0.0016 | 398 |
| rs2297638 | 6 | 131940890 | A | C | -0.0477 | 0.0067 | 0.6555 | 9.82E-13 | 88109 | 339395 | 247159 | 0.0010 | 254 |
| rs12662968 | 6 | 137288760 | A | T | 0.0558 | 0.0065 | 0.4608 | 1.09E-17 | 88109 | 339395 | 247159 | 0.0015 | 383 |
| rs7768451 | 6 | 157003689 | A | C | -0.0562 | 0.0085 | 0.8308 | 3.90E-11 | 88109 | 339395 | 247159 | 0.0009 | 220 |
| rs4709741 | 6 | 164092291 | A | C | -0.0665 | 0.011 | 0.1053 | 1.69E-09 | 88109 | 339395 | 247159 | 0.0008 | 206 |
| rs12523813 | 6 | 166223997 | T | C | -0.07 | 0.0127 | 0.9216 | 3.29E-08 | 87912 | 335059 | 246405 | 0.0007 | 175 |
| rs17168486 | 7 | 14898282 | T | C | 0.0627 | 0.0065 | 0.4194 | 4.57E-22 | 87912 | 335059 | 246405 | 0.0019 | 474 |
| rs10244051 | 7 | 15063833 | T | G | -0.067 | 0.0068 | 0.3193 | 7.33E-23 | 88109 | 339395 | 247159 | 0.0020 | 483 |
| rs3735567 | 7 | 28219310 | A | G | -0.0638 | 0.0078 | 0.2243 | 1.87E-16 | 88001 | 338685 | 246784 | 0.0014 | 351 |
| rs2240404 | 7 | 30707302 | A | G | -0.0461 | 0.0083 | 0.1946 | 2.70E-08 | 88109 | 339395 | 247159 | 0.0007 | 165 |
| rs35452727 | 7 | 44266184 | T | C | 0.0555 | 0.0085 | 0.1839 | 7.84E-11 | 87950 | 337771 | 246579 | 0.0009 | 229 |
| rs7790934 | 7 | 69701965 | A | G | -0.0649 | 0.0067 | 0.6633 | 3.82E-22 | 88109 | 339395 | 247159 | 0.0019 | 466 |
| rs39204 | 7 | 89734318 | A | C | 0.0679 | 0.0077 | 0.2244 | 7.78E-19 | 88109 | 339395 | 247159 | 0.0016 | 397 |
| rs2074120 | 7 | 93107093 | A | C | 0.0427 | 0.0068 | 0.3237 | 3.49E-10 | 88109 | 339395 | 247159 | 0.0008 | 197 |
| rs75990271 | 7 | 102336979 | T | C | 0.0693 | 0.0087 | 0.8034 | 1.68E-15 | 86692 | 331491 | 243167 | 0.0015 | 376 |
| rs2233580 | 7 | 127253550 | T | C | 0.3296 | 0.0114 | 0.0885 | 8.57E-183 | 86882 | 331561 | 243602 | 0.0175 | 4409 |
| rs3824004 | 7 | 127253551 | T | G | 0.2406 | 0.0174 | 0.036 | 1.62E-43 | 87912 | 335059 | 246405 | 0.0040 | 997 |
| rs62487901 | 7 | 140436560 | A | G | 0.0745 | 0.0122 | 0.082 | 1.19E-09 | 87164 | 330076 | 243803 | 0.0008 | 207 |
| rs887609 | 7 | 156794983 | A | G | 0.0647 | 0.0082 | 0.2203 | 4.26E-15 | 88109 | 339395 | 247159 | 0.0014 | 356 |
| rs6459732 | 7 | 156921020 | T | C | -0.0627 | 0.0108 | 0.1071 | 5.80E-09 | 88109 | 339395 | 247159 | 0.0008 | 186 |
| rs28412 | 8 | 17907387 | A | G | 0.074 | 0.013 | 0.066 | 1.31E-08 | 88109 | 339395 | 247159 | 0.0007 | 167 |
| rs56825414 | 8 | 36758946 | T | C | 0.0462 | 0.0076 | 0.2436 | 9.68E-10 | 88109 | 339395 | 247159 | 0.0008 | 195 |
| rs12680217 | 8 | 37397803 | T | C | 0.0462 | 0.0065 | 0.5546 | 1.03E-12 | 88109 | 339395 | 247159 | 0.0011 | 261 |
| rs6989203 | 8 | 41523745 | A | G | -0.1035 | 0.0093 | 0.1364 | 9.33E-29 | 88109 | 339395 | 247159 | 0.0025 | 625 |
| rs185063984 | 8 | 75203220 | A | G | -0.1337 | 0.0197 | 0.9697 | 1.18E-11 | 87955 | 338283 | 246619 | 0.0011 | 260 |
| rs12679044 | 8 | 95891356 | A | C | -0.0426 | 0.007 | 0.7127 | 1.12E-09 | 88109 | 339395 | 247159 | 0.0007 | 184 |
| rs13266634 | 8 | 118184783 | T | C | -0.1258 | 0.0064 | 0.4131 | 5.51E-85 | 88109 | 339395 | 247159 | 0.0077 | 1911 |
| rs2175055 | 9 | 1033354 | T | C | 0.0396 | 0.0065 | 0.4277 | 9.89E-10 | 88109 | 339395 | 247159 | 0.0008 | 190 |
| rs4237150 | 9 | 4290085 | C | G | 0.0846 | 0.0064 | 0.4289 | 1.22E-39 | 87912 | 335059 | 246405 | 0.0035 | 870 |
| rs10965250 | 9 | 22133284 | A | G | -0.1881 | 0.0064 | 0.4351 | 6.18E-189 | 88109 | 339395 | 247159 | 0.0174 | 4375 |
| rs7030811 | 9 | 22288140 | A | T | -0.1223 | 0.0144 | 0.0577 | 2.15E-17 | 87912 | 335059 | 246405 | 0.0016 | 403 |
| rs34532817 | 9 | 81914318 | T | C | -0.0931 | 0.0141 | 0.0556 | 4.29E-11 | 88109 | 339395 | 247159 | 0.0009 | 225 |
| rs10125947 | 9 | 83974536 | A | G | 0.0521 | 0.0067 | 0.6301 | 9.57E-15 | 88109 | 339395 | 247159 | 0.0013 | 313 |
| rs2796441 | 9 | 84308948 | A | G | -0.0792 | 0.0065 | 0.6085 | 1.01E-33 | 88109 | 339395 | 247159 | 0.0030 | 741 |
| rs113154802 | 9 | 98278413 | T | C | -0.069 | 0.0103 | 0.1154 | 1.73E-11 | 88109 | 339395 | 247159 | 0.0010 | 240 |
| rs28378473 | 9 | 139245460 | T | C | -0.1452 | 0.0119 | 0.1091 | 3.16E-34 | 84524 | 333088 | 239057 | 0.0041 | 1017 |
| rs11257655 | 10 | 12307894 | T | C | 0.1257 | 0.0065 | 0.4786 | 1.96E-84 | 88109 | 339395 | 247159 | 0.0079 | 1965 |
| rs57219676 | 10 | 12399515 | A | G | 0.0706 | 0.0119 | 0.0842 | 3.54E-09 | 88109 | 339395 | 247159 | 0.0008 | 190 |
| rs2815657 | 10 | 12548710 | A | G | 0.0423 | 0.0069 | 0.3348 | 6.92E-10 | 88109 | 339395 | 247159 | 0.0008 | 197 |
| rs117414485 | 10 | 23578177 | T | C | 0.1088 | 0.0151 | 0.047 | 5.86E-13 | 88109 | 339395 | 247159 | 0.0011 | 262 |
| rs77757273 | 10 | 63716319 | A | G | -0.0629 | 0.0095 | 0.1336 | 2.90E-11 | 88109 | 339395 | 247159 | 0.0009 | 227 |
| rs59050225 | 10 | 64950110 | T | C | -0.065 | 0.0078 | 0.2111 | 1.18E-16 | 88109 | 339395 | 247159 | 0.0014 | 348 |
| rs10824316 | 10 | 77295843 | T | C | -0.0432 | 0.0064 | 0.5164 | 1.01E-11 | 88109 | 339395 | 247159 | 0.0009 | 231 |
| rs703977 | 10 | 80944230 | T | G | 0.0614 | 0.0065 | 0.566 | 2.69E-21 | 85780 | 337056 | 242491 | 0.0019 | 459 |
| rs10887774 | 10 | 89766043 | A | G | 0.0412 | 0.0064 | 0.4636 | 9.36E-11 | 88109 | 339395 | 247159 | 0.0008 | 209 |
| rs11598977 | 10 | 94146580 | A | T | -0.1295 | 0.0148 | 0.9387 | 1.95E-18 | 88109 | 339395 | 247159 | 0.0019 | 478 |
| rs10882100 | 10 | 94460687 | C | G | 0.1418 | 0.0073 | 0.25 | 4.78E-84 | 88109 | 339395 | 247159 | 0.0075 | 1878 |
| rs10882890 | 10 | 99048262 | A | G | -0.0499 | 0.007 | 0.7083 | 1.33E-12 | 88109 | 339395 | 247159 | 0.0010 | 255 |
| rs13306146 | 10 | 112840171 | A | G | 0.0462 | 0.007 | 0.6949 | 5.06E-11 | 88109 | 339395 | 247159 | 0.0009 | 224 |
| rs7901695 | 10 | 114754088 | T | C | -0.2911 | 0.0162 | 0.9591 | 5.27E-72 | 87117 | 337721 | 244667 | 0.0066 | 1654 |
| rs11199805 | 10 | 122919206 | T | C | -0.0605 | 0.007 | 0.2941 | 7.42E-18 | 88109 | 339395 | 247159 | 0.0015 | 376 |
| rs11200595 | 10 | 124142629 | T | C | 0.0427 | 0.0066 | 0.3969 | 8.25E-11 | 88109 | 339395 | 247159 | 0.0009 | 216 |
| rs7482891 | 11 | 2197112 | A | G | 0.1107 | 0.0119 | 0.0809 | 1.60E-20 | 85516 | 334762 | 241548 | 0.0018 | 451 |
| rs7121454 | 11 | 2624456 | T | G | 0.0998 | 0.0107 | 0.8905 | 1.60E-20 | 88109 | 339395 | 247159 | 0.0019 | 481 |
| rs74728949 | 11 | 2758546 | T | C | -0.0603 | 0.0089 | 0.1592 | 1.57E-11 | 85780 | 337056 | 242491 | 0.0010 | 241 |
| rs2237897 | 11 | 2858546 | T | C | -0.2562 | 0.007 | 0.3654 | 7.13E-295 | 85780 | 337056 | 242491 | 0.0304 | 7760 |
| rs10832835 | 11 | 2880009 | A | C | 0.0576 | 0.0083 | 0.1964 | 5.01E-12 | 85583 | 332720 | 241737 | 0.0010 | 259 |
| rs2074314 | 11 | 17411821 | T | C | -0.0786 | 0.0065 | 0.6244 | 2.48E-33 | 88109 | 339395 | 247159 | 0.0029 | 718 |
| rs11030104 | 11 | 27684517 | A | G | 0.042 | 0.0064 | 0.5592 | 5.11E-11 | 88109 | 339395 | 247159 | 0.0009 | 215 |
| rs4562816 | 11 | 51247632 | A | T | 0.0387 | 0.0068 | 0.4876 | 1.01E-08 | 85193 | 330127 | 240411 | 0.0007 | 185 |
| rs628993 | 11 | 61539691 | A | G | 0.0382 | 0.0069 | 0.5065 | 2.81E-08 | 87737 | 336602 | 245867 | 0.0007 | 180 |
| rs602652 | 11 | 69462642 | A | G | 0.0509 | 0.0085 | 0.7978 | 1.83E-09 | 85780 | 337056 | 242491 | 0.0008 | 207 |
| rs11603349 | 11 | 72460694 | T | C | 0.1461 | 0.0143 | 0.9385 | 1.64E-24 | 88109 | 339395 | 247159 | 0.0025 | 610 |
| rs4331050 | 11 | 92696014 | T | G | 0.0561 | 0.009 | 0.4357 | 4.51E-10 | 42726 | 207363 | 112063 | 0.0015 | 383 |
| rs10750397 | 11 | 128234144 | A | G | 0.0401 | 0.0072 | 0.2653 | 3.13E-08 | 88109 | 339395 | 247159 | 0.0006 | 155 |
| rs4140660 | 12 | 4310220 | T | C | 0.0489 | 0.008 | 0.1998 | 1.11E-09 | 88109 | 339395 | 247159 | 0.0008 | 189 |
| rs3812821 | 12 | 4382324 | C | G | -0.0532 | 0.0071 | 0.5142 | 6.29E-14 | 87005 | 328452 | 243224 | 0.0014 | 350 |
| rs12578595 | 12 | 27964996 | T | C | -0.0679 | 0.0068 | 0.3338 | 1.43E-23 | 88109 | 339395 | 247159 | 0.0021 | 508 |
| rs80234489 | 12 | 31441179 | A | C | -0.1227 | 0.0085 | 0.8256 | 3.17E-47 | 88109 | 339395 | 247159 | 0.0043 | 1076 |
| rs7132908 | 12 | 50263148 | A | G | 0.043 | 0.0073 | 0.2573 | 3.97E-09 | 88109 | 339395 | 247159 | 0.0007 | 175 |
| rs2298640 | 12 | 50304574 | A | G | -0.0381 | 0.007 | 0.3181 | 4.80E-08 | 88109 | 339395 | 247159 | 0.0006 | 156 |
| rs2612059 | 12 | 66239111 | A | C | -0.0636 | 0.0067 | 0.6599 | 1.87E-21 | 88109 | 339395 | 247159 | 0.0018 | 450 |
| rs1913195 | 12 | 71416780 | T | G | -0.0445 | 0.0072 | 0.2771 | 6.46E-10 | 88109 | 339395 | 247159 | 0.0008 | 196 |
| rs10860209 | 12 | 97850215 | A | C | -0.0405 | 0.0065 | 0.4112 | 4.60E-10 | 88109 | 339395 | 247159 | 0.0008 | 196 |
| rs1426371 | 12 | 108629780 | A | G | -0.0485 | 0.0065 | 0.5116 | 7.26E-14 | 88109 | 339395 | 247159 | 0.0012 | 291 |
| rs2074356 | 12 | 112645401 | A | G | -0.0551 | 0.0094 | 0.1841 | 3.86E-09 | 87124 | 333814 | 244475 | 0.0009 | 226 |
| rs7132351 | 12 | 118414926 | A | G | 0.0734 | 0.0076 | 0.2435 | 2.95E-22 | 88109 | 339395 | 247159 | 0.0020 | 492 |
| rs118074491 | 12 | 121363506 | A | G | -0.2049 | 0.0197 | 0.9706 | 1.92E-25 | 87912 | 335059 | 246405 | 0.0024 | 594 |
| rs9316706 | 13 | 22589883 | A | G | 0.0436 | 0.0067 | 0.3519 | 6.27E-11 | 88109 | 339395 | 247159 | 0.0009 | 214 |
| rs10467680 | 13 | 26780420 | T | C | -0.065 | 0.0065 | 0.434 | 8.53E-24 | 88109 | 339395 | 247159 | 0.0021 | 514 |
| rs12429937 | 13 | 33571753 | A | T | -0.0727 | 0.01 | 0.8772 | 2.99E-13 | 87912 | 335059 | 246405 | 0.0011 | 282 |
| rs123378 | 13 | 51088809 | A | G | -0.0541 | 0.008 | 0.8027 | 1.69E-11 | 88109 | 339395 | 247159 | 0.0009 | 229 |
| rs1215468 | 13 | 80707429 | A | G | 0.0839 | 0.0072 | 0.719 | 1.20E-31 | 88109 | 339395 | 247159 | 0.0028 | 705 |
| rs9583907 | 13 | 91939270 | T | C | -0.0867 | 0.0086 | 0.1691 | 4.27E-24 | 88109 | 339395 | 247159 | 0.0021 | 523 |
| rs12437434 | 14 | 24878370 | T | C | -0.0524 | 0.0072 | 0.2874 | 2.53E-13 | 87912 | 335059 | 246405 | 0.0011 | 278 |
| rs61975988 | 14 | 38809661 | A | G | 0.0376 | 0.0064 | 0.4601 | 3.25E-09 | 88109 | 339395 | 247159 | 0.0007 | 174 |
| rs79823890 | 14 | 52511969 | T | G | -0.0566 | 0.0096 | 0.1332 | 3.34E-09 | 88109 | 339395 | 247159 | 0.0007 | 183 |
| rs2056855 | 14 | 77297139 | T | C | 0.0391 | 0.0067 | 0.3546 | 5.22E-09 | 88109 | 339395 | 247159 | 0.0007 | 173 |
| rs72627178 | 14 | 77372210 | A | G | -0.0505 | 0.0068 | 0.6384 | 8.23E-14 | 85780 | 337056 | 242491 | 0.0012 | 291 |
| rs73347525 | 14 | 101255172 | A | G | 0.0669 | 0.0082 | 0.7579 | 3.20E-16 | 85621 | 335432 | 241911 | 0.0016 | 407 |
| rs55700915 | 14 | 103237952 | A | G | 0.0426 | 0.0066 | 0.4389 | 9.24E-11 | 87912 | 335059 | 246405 | 0.0009 | 221 |
| rs76704029 | 15 | 28546173 | T | C | 0.0464 | 0.0084 | 0.7115 | 3.58E-08 | 77339 | 249112 | 209548 | 0.0009 | 219 |
| rs8043085 | 15 | 38828140 | T | G | 0.0514 | 0.0065 | 0.4495 | 2.14E-15 | 88109 | 339395 | 247159 | 0.0013 | 324 |
| rs3743140 | 15 | 40616742 | A | G | 0.077 | 0.0078 | 0.2331 | 3.71E-23 | 85583 | 332720 | 241737 | 0.0021 | 525 |
| rs149336329 | 15 | 52587740 | T | G | -0.112 | 0.0149 | 0.056 | 5.23E-14 | 87912 | 335059 | 246405 | 0.0013 | 328 |
| rs7161785 | 15 | 62395224 | C | G | -0.0847 | 0.0065 | 0.4154 | 2.71E-39 | 88109 | 339395 | 247159 | 0.0035 | 864 |
| rs4299117 | 15 | 75789444 | T | C | -0.0531 | 0.0065 | 0.3926 | 4.69E-16 | 88109 | 339395 | 247159 | 0.0013 | 333 |
| rs8034133 | 15 | 77763712 | C | G | -0.0848 | 0.0065 | 0.6035 | 5.63E-39 | 88109 | 339395 | 247159 | 0.0034 | 854 |
| rs2290202 | 15 | 91512267 | T | G | 0.0677 | 0.0064 | 0.488 | 6.58E-26 | 88109 | 339395 | 247159 | 0.0023 | 567 |
| rs7167984 | 15 | 93832067 | A | G | -0.047 | 0.0066 | 0.5575 | 7.21E-13 | 88109 | 339395 | 247159 | 0.0011 | 270 |
| rs740862 | 16 | 3689678 | T | G | -0.0384 | 0.0066 | 0.5274 | 4.94E-09 | 88109 | 339395 | 247159 | 0.0007 | 182 |
| rs73541251 | 16 | 20331737 | C | G | -0.0987 | 0.0118 | 0.9209 | 6.84E-17 | 88109 | 339395 | 247159 | 0.0014 | 351 |
| rs11642015 | 16 | 53802494 | T | C | 0.1299 | 0.0085 | 0.1751 | 3.46E-53 | 88109 | 339395 | 247159 | 0.0049 | 1211 |
| rs12051517 | 16 | 72022866 | A | G | -0.0358 | 0.0064 | 0.5687 | 2.15E-08 | 88109 | 339395 | 247159 | 0.0006 | 155 |
| rs6416749 | 16 | 73100308 | T | C | -0.0565 | 0.0069 | 0.6104 | 1.71E-16 | 87753 | 333435 | 245826 | 0.0015 | 376 |
| rs2925979 | 16 | 81534790 | T | C | 0.0422 | 0.0066 | 0.3656 | 1.73E-10 | 88109 | 339395 | 247159 | 0.0008 | 204 |
| rs13342232 | 17 | 6945940 | A | G | -0.1196 | 0.011 | 0.9047 | 1.10E-27 | 88109 | 339395 | 247159 | 0.0025 | 611 |
| rs2905798 | 17 | 29536990 | A | G | 0.0444 | 0.0071 | 0.671 | 2.95E-10 | 88109 | 339395 | 247159 | 0.0009 | 215 |
| rs35551980 | 17 | 36049552 | T | C | -0.0726 | 0.0074 | 0.7469 | 1.02E-22 | 87912 | 335059 | 246405 | 0.0020 | 494 |
| rs11651052 | 17 | 36102381 | A | G | 0.1235 | 0.007 | 0.3058 | 2.46E-69 | 87950 | 337771 | 246579 | 0.0065 | 1611 |
| rs11650046 | 17 | 76777694 | A | C | -0.038 | 0.0065 | 0.4006 | 4.77E-09 | 88109 | 339395 | 247159 | 0.0007 | 172 |
| rs9948462 | 18 | 7076836 | T | C | 0.049 | 0.0071 | 0.7024 | 7.40E-12 | 88109 | 339395 | 247159 | 0.0010 | 248 |
| rs7235574 | 18 | 42368034 | A | G | -0.0419 | 0.0074 | 0.2552 | 1.29E-08 | 88109 | 339395 | 247159 | 0.0007 | 165 |
| rs571312 | 18 | 57839769 | A | C | 0.0887 | 0.0077 | 0.216 | 7.65E-31 | 88109 | 339395 | 247159 | 0.0027 | 660 |
| rs12957245 | 18 | 74614501 | T | C | -0.0382 | 0.0067 | 0.3402 | 1.47E-08 | 87912 | 335059 | 246405 | 0.0007 | 162 |
| rs8101064 | 19 | 7293119 | T | C | 0.0641 | 0.0104 | 0.1264 | 6.47E-10 | 87912 | 335059 | 246405 | 0.0009 | 224 |
| rs2115107 | 19 | 7968168 | A | G | 0.0374 | 0.0068 | 0.3534 | 4.54E-08 | 85780 | 337056 | 242491 | 0.0006 | 158 |
| rs150182828 | 19 | 21856746 | T | C | 0.2367 | 0.0198 | 0.9658 | 8.19E-33 | 87912 | 335059 | 246405 | 0.0037 | 918 |
| rs10422861 | 19 | 33894846 | T | C | -0.0565 | 0.0065 | 0.4636 | 2.06E-18 | 85583 | 332720 | 241737 | 0.0016 | 393 |
| rs58304657 | 19 | 46176405 | C | G | 0.0778 | 0.0065 | 0.514 | 8.29E-33 | 85621 | 335432 | 241911 | 0.0030 | 750 |
| rs6036116 | 20 | 22430350 | T | C | 0.0405 | 0.0066 | 0.6391 | 9.66E-10 | 88109 | 339395 | 247159 | 0.0008 | 187 |
| rs12625671 | 20 | 42994812 | T | C | -0.0673 | 0.0065 | 0.5594 | 2.89E-25 | 88109 | 339395 | 247159 | 0.0022 | 553 |
| rs6122888 | 20 | 48826880 | T | C | 0.0521 | 0.0065 | 0.5086 | 1.31E-15 | 88109 | 339395 | 247159 | 0.0014 | 336 |
| rs7121 | 20 | 57478807 | T | C | 0.0393 | 0.0066 | 0.5972 | 2.52E-09 | 88109 | 339395 | 247159 | 0.0007 | 184 |
| rs5762925 | 22 | 29369398 | A | C | -0.0355 | 0.0064 | 0.5065 | 3.53E-08 | 88109 | 339395 | 247159 | 0.0006 | 156 |
| rs28637892 | 22 | 46313618 | T | G | 0.0563 | 0.0081 | 0.2192 | 3.00E-12 | 80101 | 322056 | 228723 | 0.0011 | 268 |
| rs6520050 | 22 | 46478195 | A | G | 0.0596 | 0.0098 | 0.849 | 1.30E-09 | 85583 | 332720 | 241737 | 0.0009 | 225 |
| rs28645887 | 22 | 50356274 | T | C | -0.0636 | 0.007 | 0.4436 | 6.67E-20 | 85583 | 332720 | 241737 | 0.0020 | 494 |

^a^ Based on hg19, build 37.

Abbreviations: SNP, single nucleotide polymorphism; CHR, chromosome; POS, position; EAF, effective allele frequency.

### Supplementary Table 4: Baseline characteristics of participants categorised by T2DM status at follow-up^a^

| **Characteristic** | **Men** | | **Women** | |
| --- | --- | --- | --- | --- |
|  | **New-onset T2DM** | **No diabetes at follow-up** | **New-onset T2DM** | **No diabetes at follow-up** |
|  | (N=9,206) | (N=188,450) | (N=14,392) | (N=268,090) |
| Baseline age, years (mean, SD) | 55.0(10.2) | 52.4(10.9) | 55.3(9.5) | 50.7(10.4) |
| Age at T2DM diagnosis (mean, SD) | 62.4(10.2) | .. | 62.6(9.6) | .. |
| **Socioeconomic and lifestyle factors** |  | | | |
| ≥ 6 years of education, N (%) | 5,100(55.4) | 108,560(57.6) | 4,742(32.9) | 118,796(44.3) |
| Annual household income ≥ 35,000 yuan, N (%) | 2,367(25.7) | 37,243(19.8) | 2,723(18.9) | 43,768(16.3) |
| Ever regular smokers^b^, N (%) | 6,874(74.7) | 140,374(74.5) | 716(5.0) | 8,083(3.0) |
| Ever regular alcohol drinkers^c^, N (%) | 3,578(38.9) | 69,664(37.0) | 386(2.7) | 6,763(2.5) |
| Physical activity, MET h/d (mean, SD) | 20.5(14.9) | 22.5(15.3) | 19.1(12.7) | 20.9(12.8) |
| **Anthropometry** |  | | | |
| BMI, kg/m^2^ (mean, SD) | 25.5(3.4) | 23.3(3.2) | 25.8(3.7) | 23.6(3.4) |
| General obese^d^, N (%) | 2,033(22.1) | 14,660(7.8) | 3,725(25.9) | 2,774(10.3) |
| WC, cm (mean, SD) | 87.9(10.0) | 81.4(9.6) | 84.4(9.8) | 78.4(9.3) |
| Central obese^e^, N (%) | 4,015(43.6) | 37,323(19.8) | 6,833(47.5) | 61,988(23.1) |
| **Disease history** |  | | | |
| Cirrhosis/chronic hepatitis, N (%) | 187(2.0) | 3,197(1.7) | 139(1.0) | 2,225(0.8) |

Abbreviations: MET, metabolic equivalent task; BMI, body mass index; WC, waist circumference.

^a^ Regardless of cancer diagnosis. ^b^ Ever regular smokers included former and current regular smokers. ^c^ Ever regular alcohol drinkers included former regular and current weekly drinkers. ^d^ General obesity was defined by a BMI ≥ 28 kg/m^2^. ^e^ Central obesity was defined by a WC ≥ 90 cm in men and ≥ 85 cm in women, respectively.

### Supplementary Table 5: Characteristics in participants with new-onset T2DM who failed to find any match in 1:3 matching

| **Men (N=37)** | | | | **Women (N=36)** | | | |
| --- | --- | --- | --- | --- | --- | --- | --- |
| **Strata number^a^** | **Study region** | **Baseline body mass index, kg/m^2^** | **Age at T2DM diagnosis, years** | **Strata number^a^** | **Study region** | **Baseline body mass index, kg/m^2^** | **Age at T2DM diagnosis, years** |
| 8375 | *Zhejiang* | 28.6 | 78.9 | 9010 | *Zhejiang* | 30.1 | 78.8 |
| 8685 | *Zhejiang* | 28.9 | 76.4 | 5865 | *Zhejiang* | 31.4 | 78.5 |
| 8611 | *Zhejiang* | 29.9 | 79.3 | 8834 | *Sichuan* | 33.3 | 87.1 |
| 6491 | *Zhejiang* | 30.5 | 74.2 | 5315 | *Zhejiang* | 33.9 | 72.5 |
| 8141 | *Zhejiang* | 30.6 | 69.4 | 11441 | *Sichuan* | 34.3 | 62.8 |
| 8295 | *Zhejiang* | 30.9 | 73.1 | 12463 | *Hunan* | 34.4 | 83.7 |
| 5904 | *Sichuan* | 31.5 | 69.7 | 10237 | *Henan* | 34.6 | 85.4 |
| 8430 | *Sichuan* | 31.6 | 69.6 | 11231 | *Sichuan* | 35.0 | 78.5 |
| 6159 | *Zhejiang* | 31.9 | 64.1 | 401 | *Zhejiang* | 35.2 | 62.7 |
| 7659 | *Sichuan* | 32.0 | 64.4 | 8288 | *Sichuan* | 36.1 | 52.9 |
| 7906 | *Harbin* | 32.9 | 74.0 | 1220 | *Sichuan* | 36.3 | 72.3 |
| 75 | *Zhejiang* | 33.0 | 75.1 | 10713 | *Sichuan* | 36.4 | 70.7 |
| 2513 | *Gansu* | 33.0 | 73.0 | 1431 | *Zhejiang* | 36.6 | 57.6 |
| 7271 | *Liuzhou* | 33.4 | 52.0 | 12776 | *Haikou* | 36.8 | 80.3 |
| 7599 | *Haikou* | 33.4 | 72.4 | 2758 | *Haikou* | 37.2 | 66.4 |
| 1603 | *Gansu* | 33.9 | 47.8 | 12729 | *Haikou* | 37.7 | 46.0 |
| 2194 | *Gansu* | 34.1 | 61.7 | 4084 | *Harbin* | 37.8 | 83.0 |
| 1955 | *Liuzhou* | 34.3 | 80.2 | 6020 | *Zhejiang* | 38.2 | 64.7 |
| 2021 | *Sichuan* | 35.0 | 59.8 | 13198 | *Gansu* | 38.3 | 54.1 |
| 8537 | *Harbin* | 35.2 | 62.2 | 8434 | *Sichuan* | 38.6 | 70.4 |
| 5781 | *Qingdao* | 35.6 | 57.3 | 2433 | *Haikou* | 38.9 | 59.1 |
| 5799 | *Harbin* | 36.0 | 71.0 | 3596 | *Harbin* | 39.2 | 53.3 |
| 3785 | *Harbin* | 36.4 | 69.3 | 11645 | *Hunan* | 39.2 | 50.4 |
| 7190 | *Suzhou* | 36.7 | 53.2 | 12522 | *Henan* | 39.7 | 60.3 |
| 6569 | *Zhejiang* | 37.3 | 70.0 | 3589 | *Harbin* | 40.1 | 62.1 |
| 4622 | *Sichuan* | 37.6 | 72.9 | 4265 | *Sichuan* | 40.2 | 49.2 |
| 5591 | *Henan* | 37.9 | 50.5 | 13051 | *Gansu* | 40.4 | 52.1 |
| 3057 | *Zhejiang* | 38.2 | 50.0 | 8804 | *Suzhou* | 40.6 | 69.7 |
| 6049 | *Sichuan* | 38.2 | 46.8 | 2341 | *Liuzhou* | 41.3 | 52.7 |
| 789 | *Qingdao* | 38.5 | 58.1 | 10319 | *Harbin* | 41.3 | 66.9 |
| 1842 | *Harbin* | 38.8 | 48.7 | 3790 | *Sichuan* | 41.8 | 47.0 |
| 5394 | *Haikou* | 39.3 | 60.4 | 2931 | *Haikou* | 41.9 | 61.1 |
| 3718 | *Zhejiang* | 39.6 | 61.9 | 9462 | *Harbin* | 42.0 | 57.5 |
| 454 | *Liuzhou* | 41.8 | 61.5 | 8363 | *Henan* | 43.3 | 79.2 |
| 6525 | *Sichuan* | 43.3 | 51.6 | 8053 | *Harbin* | 46.2 | 65.4 |
| 8391 | *Henan* | 45.7 | 48.9 | 1666 | *Suzhou* | 51.0 | 58.4 |
| 2962 | *Harbin* | 47.9 | 62.9 | .. | .. | .. | .. |

^a^ Strata number was assigned separately for men and women based on the calendar time of their T2DM diagnosis, following a chronologic order.

### Supplementary Table 6: Examination in censoring patterns following 1:3 exact matching

| **Event/censoring** | **Men** | | **Women** | |
| --- | --- | --- | --- | --- |
|  | **New-onset T2DM** | **Matched unexposed** | **New-onset T2DM** | **Matched unexposed** |
|  | (N=8,657) | (N=25,852) | (N=13,680) | (N=40,938) |
| Incident cancer diagnosis, N % | 542(6.3) | 964(3.7) | 582(4.3) | 1262(3.1) |
| Administrative censoring, N % | 7230(83.5) | 22191(85.8) | 12120(88.6) | 36170(88.4) |
| Loss to follow-up, N % | 4(0.05) | 123(0.5) | 13(0.1) | 199(0.5) |
| Non-cancer death^a^, N % | 881(10.2) | 1366(5.3) | 965(7.1) | 1321(3.2) |
| Incident T2DM diagnosis^b^, N % | .. | 1208(4.7) | .. | 1986(4.9) |

^a^ Before a cancer or T2DM diagnosis (in matched controls). ^b^ Before a cancer diagnosis.

### Supplementary Table 7: Comparison of effect estimates from cause-specific and subdistribution hazard models for T2DM on first primary cancer incidence

| **Outcome** | **Men** | | **Women** | |
| --- | --- | --- | --- | --- |
|  | **Subdistribution hazard model** | **Cause-specific hazard model** | **Subdistribution hazard model** | **Cause-specific hazard model** |
|  | **HR (95% CI)** | **HR (95% CI)** | **HR (95% CI)** | **HR (95% CI)** |
| Total first primary cancers | 1.49(1.32–1.69) | 1.57(1.38–1.78) | 1.18(1.06–1.32) | 1.29(1.14–1.46) |
| ORCs | 1.64(1.31–2.04) | 1.73(1.37–2.19) | 1.12(0.94–1.34) | 1.34(1.10–1.62) |
| Total cancers minus ORCs | 1.42(1.22–1.64) | 1.49(1.28–1.74) | 1.22(1.06–1.41) | 1.27(1.09–1.47) |
| Digestive tract organ cancers | 1.27(1.02–1.57) | 1.33(1.06–1.67) | 1.11(0.87–1.41) | 1.14(0.89–1.46) |
| Digestive accessory organ cancers | 2.12(1.56–2.89) | 2.23(1.59–3.11) | 1.79(1.26–2.53) | 2.94(1.94–4.45) |
| ORCs in reproductive organs | .. | .. | 0.88(0.67–1.15) | 0.90(0.68–1.20) |
| Site-specific cancer |  | | | |
| Liver | 2.03(1.40–2.93) | 2.12(1.42–3.15) | 1.64(1.06–2.53) | 2.39(1.42–4.04) |
| Pancreas | 2.34(1.32–4.14) | 2.57(1.35–4.92) | 2.21(1.25–3.89) | 3.95(1.92–8.13) |
| Colorectum | 1.29(0.92–1.81) | 1.33(0.93–1.91) | 1.11(0.79–1.55) | 1.16(0.82–1.64) |
| Lung | 1.45(1.11–1.89) | 1.55(1.17–2.05) | 1.12(0.85–1.46) | 1.15(0.87–1.52) |
| Stomach | 1.08(0.75–1.56) | 1.13(0.77–1.68) | 1.11(0.72–1.69) | 1.11(0.72–1.71) |
| Breast (overall) | .. | .. | 0.98(0.73–1.33) | 1.01(0.73–1.38) |
| Postmenopausal breast | .. | .. | 0.96(0.70–1.31) | 1.02(0.74–1.41) |

Abbreviations: ORCs, obesity-related cancers; HR, hazard ratio.

Each cell contains the HR and associated 95% CI for the given outcome and for the follow-up time window beyond 12 months in new-onset T2DM compared to the matched unexposed following 1-3 exact matching. All models were stratified on the matched set and further adjusted for waist circumference, smoking status, alcohol consumption habit, MET hours, highest education level, annual household income, and cirrhosis/chronic hepatitis history (for liver cancer and for composite cancer outcomes that encompassed liver cancer).

### Supplementary Table 8: Associations of new-onset T2DM with composite cancer risks after applying a narrower caliper width in BMI matching

|  | | **Men** | | **Women** | |
| --- | --- | --- | --- | --- | --- |
|  |  | **New-onset T2DM** | **Matched unexposed** | **New-onset T2DM** | **Matched unexposed** |
| No. of participants | | 8,610 | 25,627 | 13,642 | 40,700 |
| No. of person-years | | 39,493 | 119,350 | 67,123 | 199,269 |
| *Total first primary cancers* | |  |  |  |  |
| No. of cases | | 539 | 1,001 | 580 | 1,243 |
| IR^a^ | | 1364.8(1249.6–1480.0) | 838.7(786.8–890.7) | 864.1(793.8–934.4) | 623.8(589.1–658.5) |
| HR (95% CI)^b^ | Model 1^c^ | 1.43(1.26–1.62) | 1(ref) | 1.33(1.19–1.50) | 1(ref) |
|  | Model 2^d^ | 1.40(1.24–1.59) | 1(ref) | 1.33(1.18–1.49) | 1(ref) |
| *ORCs* | |  |  |  |  |
| No. of cases | | 175 | 284 | 220 | 475 |
| IR^a^ | | 443.1(377.5–508.8) | 238.0(210.3–265.6) | 327.8(284.4–371.1) | 238.4(216.9–259.8) |
| HR (95% CI)^b^ | Model 1^c^ | 1.64(1.31–2.06) | 1(ref) | 1.44(1.18–1.74) | 1(ref) |
|  | Model 2^d^ | 1.60(1.27–2.02) | 1(ref) | 1.44(1.18–1.75) | 1(ref) |
| *Total cancers minus ORCs* | |  |  |  |  |
| No. of cases | | 364 | 717 | 360 | 768 |
| IR^a^ | | 921.7(827.0–1016.4) | 600.8(556.8–644.7) | 536.3(480.9–591.7) | 385.4(358.2–412.7) |
| HR (95% CI)^b^ | Model 1^c^ | 1.34(1.16–1.56) | 1(ref) | 1.33(1.15–1.54) | 1(ref) |
|  | Model 2^d^ | 1.33(1.14–1.55) | 1(ref) | 1.32(1.14–1.53) | 1(ref) |
| *Digestive tract organ cancers* | |  |  |  |  |
| No. of cases | | 160 | 353 | 127 | 327 |
| IR^a^ | | 405.1(342.4–467.9) | 295.8(264.9–326.6) | 189.2(156.3–222.1) | 164.1(146.3–181.9) |
| HR (95% CI)^b^ | Model 1^c^ | 1.22(0.98–1.53) | 1(ref) | 1.08(0.85–1.38) | 1(ref) |
|  | Model 2^d^ | 1.23(0.98–1.54) | 1(ref) | 1.07(0.84–1.36) | 1(ref) |
| *Digestive accessory organ cancers* | |  |  |  |  |
| No. of cases | | 103 | 134 | 74 | 91 |
| IR^a^ | | 260.8(210.4–311.2) | 112.3(93.3–131.3) | 110.2(85.1–135.4) | 45.7(36.3–55.0) |
| HR (95% CI)^b^ | Model 1^c^ | 1.94(1.42–2.65) | 1(ref) | 2.43(1.67–3.55) | 1(ref) |
|  | Model 2^d^ | 1.87(1.35–2.58) | 1(ref) | 2.45(1.66–3.60) | 1(ref) |
| *ORCs in reproductive organs* | |  |  |  |  |
| No. of cases | | .. | .. | 78 | 223 |
| IR^a^ | | .. | .. | 116.2(90.4–142.0) | 111.9(97.2–126.6) |
| HR (95% CI)^b^ | Model 1^c^ | .. | .. | 1.09(0.82–1.46) | 1(ref) |
|  | Model 2^d^ | .. | .. | 1.12(0.84–1.50) | 1(ref) |

Abbreviations: ORCs, obesity-related cancers; IR, incidence rate; HR, hazard ratio.

^a^ Per 100,000 person-years. ^b^ The HR for follow-up time window over 12 months was presented. ^c^ Model 1 was stratified on the matched set. ^d^ Model 2 was stratified on the matched set and further adjusted for waist circumference, smoking status, alcohol consumption habit, physical activity, highest education level, annual household income, and cirrhosis/chronic hepatitis history (for composite cancer risks that encompassed liver cancer).

### Supplementary Table 9: Associations of new-onset T2DM with site-specific cancer risks after applying a narrower caliper width in BMI matching

|  | | **Men** | | **Women** | |
| --- | --- | --- | --- | --- | --- |
|  |  | **New-onset T2DM** | **Matched unexposed** | **New-onset T2DM** | **Matched unexposed** |
| No. of participants | | 8,610 | 25,627 | 13,642 | 40,700 |
| No. of person-years | | 39,493 | 119,350 | 67,123 | 199,269 |
| *Liver* | |  |  |  |  |
| No. of cases | | 67 | 98 | 42 | 52 |
| IR^a^ | | 169.7(129.0–210.3) | 82.1(65.9–98.4) | 62.6(43.6–81.5) | 26.1(19.0–33.2) |
| HR (95% CI)^b^ | Model 1^c^ | 1.83(1.27–2.64) | 1(ref) | 2.32(1.44–3.71) | 1(ref) |
|  | Model 2^d^ | 1.69(1.15–2.50) | 1(ref) | 2.34(1.44–3.81) | 1(ref) |
| *Pancreas* | |  |  |  |  |
| No. of cases | | 36 | 36 | 33 | 39 |
| IR^a^ | | 91.2(61.4–120.9) | 30.2(20.3–40.0) | 49.2(32.4–65.9) | 19.6(13.4–25.7) |
| HR (95% CI)^b^ | Model 1^c^ | 2.27(1.27–4.07) | 1(ref) | 1.88(1.08–3.28) | 1(ref) |
|  | Model 2^d^ | 2.50(1.33–4.72) | 1(ref) | 1.84(1.02–3.30) | 1(ref) |
| *Colorectum* | |  |  |  |  |
| No. of cases | | 62 | 139 | 64 | 151 |
| IR^a^ | | 157.0(117.9–196.1) | 116.5(97.1–135.8) | 95.3(72.0–118.7) | 75.8(63.7–87.9) |
| HR (95% CI)^b^ | Model 1^c^ | 1.35(0.96–1.91) | 1(ref) | 1.21(0.86–1.71) | 1(ref) |
|  | Model 2^d^ | 1.34(0.94–1.90) | 1(ref) | 1.21(0.85–1.71) | 1(ref) |
| *Lung* | |  |  |  |  |
| No. of cases | | 115 | 236 | 101 | 211 |
| IR^a^ | | 291.2(238.0–344.4) | 197.7(172.5–223.0) | 150.5(121.1–179.8) | 105.9(91.6–120.2) |
| HR (95% CI)^b^ | Model 1^c^ | 1.30(1.00–1.70) | 1(ref) | 1.30(0.98–1.72) | 1(ref) |
|  | Model 2^d^ | 1.33(1.01–1.75) | 1(ref) | 1.27(0.95–1.68) | 1(ref) |
| *Stomach* | |  |  |  |  |
| No. of cases | | 59 | 132 | 40 | 106 |
| IR^a^ | | 149.4(111.3–187.5) | 110.6(91.7–129.5) | 59.6(41.1–78.1) | 53.2(43.1–63.3) |
| HR (95% CI)^b^ | Model 1^c^ | 0.99(0.68–1.46) | 1(ref) | 0.92(0.61–1.39) | 1(ref) |
|  | Model 2^d^ | 1.02(0.69–1.51) | 1(ref) | 0.92(0.61–1.40) | 1(ref) |
| *Breast (overall)* | |  |  |  |  |
| No. of cases | | .. | .. | 66 | 164 |
| IR^a^ | | .. | .. | 98.3(74.6–122.0) | 82.3(69.7–94.9) |
| HR (95% CI)^b^ | Model 1^c^ | .. | .. | 1.23(0.89–1.69) | 1(ref) |
|  | Model 2^d^ | .. | .. | 1.26(0.91–1.74) | 1(ref) |
| *Postmenopausal breast* | |  |  |  |  |
| No. of cases | | .. | .. | 61 | 153 |
| IR^a^ | | .. | .. | 90.9(68.1–113.7) | 76.8(64.6–88.9) |
| HR (95% CI)^b^ | Model 1^c^ | .. | .. | 1.23(0.89–1.72) | 1(ref) |
|  | Model 2^d^ | .. | .. | 1.27(0.91–1.77) | 1(ref) |

Abbreviations: IR, incidence rate; HR, hazard ratio.

^a^ Per 100,000 person-years. ^b^ The HR for follow-up time window over 12 months was presented. ^c^ Model 1 was stratified on the matched set. ^d^ Model 2 was stratified on the matched set and further adjusted for waist circumference, smoking status, alcohol consumption habit, physical activity, highest education level, annual household income, and cirrhosis/chronic hepatitis history (for liver cancer).

### Supplementary Table 10: Associations of new-onset T2DM with composite cancer risks following 1:5 exact matching

|  | | **Men** | | **Women** | |
| --- | --- | --- | --- | --- | --- |
|  |  | **New-onset T2DM** | **Matched unexposed** | **New-onset T2DM** | **Matched unexposed** |
| No. of participants | | 8,568 | 42,326 | 13,586 | 67,365 |
| No. of person-years | | 39,539 | 199,595 | 67,108 | 331,540 |
| *Total first primary cancers* | |  |  |  |  |
| No. of cases | | 540 | 1657 | 577 | 2076 |
| IR^a^ | | 1365.7(1250.5–1480.9) | 830.2(790.2–870.2) | 859.8(789.7–930.0) | 626.2(599.2–653.1) |
| HR (95% CI)^b^ | Model 1^c^ | 1.52(1.35–1.70) | 1(ref) | 1.28(1.16–1.43) | 1(ref) |
|  | Model 2^d^ | 1.50(1.34–1.69) | 1(ref) | 1.27(1.14–1.42) | 1(ref) |
| *ORCs* | |  |  |  |  |
| No. of cases | | 176 | 441 | 219 | 846 |
| IR^a^ | | 445.1(379.4–510.9) | 220.9(200.3–241.6) | 326.3(283.1–369.6) | 255.2(238.0–272.4) |
| HR (95% CI)^b^ | Model 1^c^ | 1.78(1.44–2.19) | 1(ref) | 1.36(1.13–1.62) | 1(ref) |
|  | Model 2^d^ | 1.76(1.42–2.17) | 1(ref) | 1.34(1.12–1.60) | 1(ref) |
| *Total cancers minus ORCs* | |  |  |  |  |
| No. of cases | | 364 | 1216 | 358 | 1230 |
| IR^a^ | | 920.6(826.0–1015.2) | 609.2(575.0–643.5) | 533.5(478.2–588.7) | 371.0(350.3–391.7) |
| HR (95% CI)^b^ | Model 1^c^ | 1.42(1.23–1.63) | 1(ref) | 1.34(1.17–1.54) | 1(ref) |
|  | Model 2^d^ | 1.41(1.22–1.62) | 1(ref) | 1.33(1.16–1.52) | 1(ref) |
| *Digestive tract organ cancers* | |  |  |  |  |
| No. of cases | | 161 | 599 | 124 | 518 |
| IR^a^ | | 407.2(344.3–470.1) | 300.1(276.1–324.1) | 184.8(152.3–217.3) | 156.2(142.8–169.7) |
| HR (95% CI)^b^ | Model 1^c^ | 1.29(1.05–1.58) | 1(ref) | 1.12(0.89–1.41) | 1(ref) |
|  | Model 2^d^ | 1.31(1.07–1.61) | 1(ref) | 1.10(0.87–1.38) | 1(ref) |
| *Digestive accessory organ cancers* | |  |  |  |  |
| No. of cases | | 104 | 183 | 74 | 172 |
| IR^a^ | | 263.0(212.5–313.6) | 91.7(78.4–105.0) | 110.3(85.1–135.4) | 51.9(44.1–59.6) |
| HR (95% CI)^b^ | Model 1^c^ | 2.60(1.94–3.49) | 1(ref) | 2.11(1.51–2.95) | 1(ref) |
|  | Model 2^d^ | 2.46(1.81–3.35) | 1(ref) | 2.12(1.51–2.97) | 1(ref) |
| *ORCs in reproductive organs* | |  |  |  |  |
| No. of cases | | .. | .. | 79 | 387 |
| IR^a^ | | .. | .. | 117.7(91.8–143.7) | 116.7(105.1–128.4) |
| HR (95% CI)^b^ | Model 1^c^ | .. | .. | 1.04(0.79–1.35) | 1(ref) |
|  | Model 2^d^ | .. | .. | 1.03(0.79–1.35) | 1(ref) |

Abbreviations: ORCs, obesity-related cancers; IR, incidence rate; HR, hazard ratio.

^a^ Per 100,000 person-years. ^b^ The HR for follow-up time window over 12 months was presented. ^c^ Model 1 was stratified on the matched set. ^d^ Model 2 was stratified on the matched set and further adjusted for waist circumference, smoking status, alcohol consumption habit, physical activity, highest education level, annual household income, and cirrhosis/chronic hepatitis history (for composite cancer risks that encompassed liver cancer).

### Supplementary Table 11: Associations of new-onset T2DM with site-specific cancer risks following 1:5 exact matching

|  | | **Men** | | **Women** | |
| --- | --- | --- | --- | --- | --- |
|  |  | **New-onset T2DM** | **Matched unexposed** | **New-onset T2DM** | **Matched unexposed** |
| No. of participants | | 8,568 | 42,326 | 13,586 | 67,365 |
| No. of person-years | | 39,539 | 199,595 | 67,108 | 331,540 |
| *Liver* | |  |  |  |  |
| No. of cases | | 68 | 137 | 42 | 125 |
| IR^a^ | | 172.0(131.1–212.9) | 68.6(57.1–80.1) | 62.6(43.7–81.5) | 37.7(31.1–44.3) |
| HR (95% CI)^b^ | Model 1^c^ | 2.47(1.75–3.50) | 1(ref) | 1.83(1.21–2.78) | 1(ref) |
|  | Model 2^d^ | 2.32(1.61–3.34) | 1(ref) | 1.85(1.21–2.83) | 1(ref) |
| *Pancreas* | |  |  |  |  |
| No. of cases | | 36 | 47 | 33 | 48 |
| IR^a^ | | 91.0(61.3–120.8) | 23.5(16.8–30.3) | 49.2(32.4–66.0) | 14.5(10.4–18.6) |
| HR (95% CI)^b^ | Model 1^c^ | 2.99(1.71–5.23) | 1(ref) | 2.44(1.43–4.19) | 1(ref) |
|  | Model 2^d^ | 3.71(2.03–6.81) | 1(ref) | 2.45(1.41–4.25) | 1(ref) |
| *Colorectum* | |  |  |  |  |
| No. of cases | | 62 | 226 | 62 | 267 |
| IR^a^ | | 156.8(117.8–195.8) | 113.2(98.5–128.0) | 92.4(69.4–115.4) | 80.5(70.9–90.2) |
| HR (95% CI)^b^ | Model 1^c^ | 1.32(0.96–1.82) | 1(ref) | 1.06(0.77–1.45) | 1(ref) |
|  | Model 2^d^ | 1.31(0.95–1.80) | 1(ref) | 1.03(0.75–1.42) | 1(ref) |
| *Lung* | |  |  |  |  |
| No. of cases | | 115 | 348 | 101 | 338 |
| IR^a^ | | 290.9(237.7–344.0) | 174.4(156.0–192.7) | 150.5(121.2–179.9) | 101.9(91.1–112.8) |
| HR (95% CI)^b^ | Model 1^c^ | 1.53(1.19–1.97) | 1(ref) | 1.37(1.06–1.77) | 1(ref) |
|  | Model 2^d^ | 1.52(1.17–1.96) | 1(ref) | 1.33(1.02–1.73) | 1(ref) |
| *Stomach* | |  |  |  |  |
| No. of cases | | 60 | 224 | 39 | 151 |
| IR^a^ | | 151.7(113.4–190.1) | 112.2(97.5–126.9) | 58.1(39.9–76.4) | 45.5(38.3–52.8) |
| HR (95% CI)^b^ | Model 1^c^ | 1.13(0.79–1.62) | 1(ref) | 1.32(0.87–2.00) | 1(ref) |
|  | Model 2^d^ | 1.19(0.83–1.71) | 1(ref) | 1.30(0.86–1.98) | 1(ref) |
| *Breast (overall)* | |  |  |  |  |
| No. of cases | | .. | .. | 68 | 301 |
| IR^a^ | | .. | .. | 101.3(77.2–125.4) | 90.8(80.5–101.0) |
| HR (95% CI)^b^ | Model 1^c^ | .. | .. | 1.11(0.83–1.49) | 1(ref) |
|  | Model 2^d^ | .. | .. | 1.12(0.83–1.50) | 1(ref) |
| *Postmenopausal breast* | |  |  |  |  |
| No. of cases | | .. | .. | 62 | 286 |
| IR^a^ | | .. | .. | 92.4(69.4–115.4) | 86.3(76.3–96.3) |
| HR (95% CI)^b^ | Model 1^c^ | .. | .. | 1.09(0.81–1.48) | 1(ref) |
|  | Model 2^d^ | .. | .. | 1.10(0.81–1.49) | 1(ref) |

Abbreviations: IR, incidence rate; HR, hazard ratio.

a Per 100,000 person-years. b The HR for follow-up time window over 12 months was presented. c Model 1 was stratified on the matched set. d Model 2 was stratified on the matched set and further adjusted for waist circumference, smoking status, alcohol consumption habit, physical activity, highest education level, annual household income, and cirrhosis/chronic hepatitis history (for liver cancer).

### Supplementary Table 12: Associations of new-onset T2DM with composite cancer risks in smoking- and alcohol-naïve women

|  | | **Smoking- and alcohol-naïve women** | |
| --- | --- | --- | --- |
|  |  | **New-onset T2DM** | **Matched unexposed** |
| No. of participants | | 9,052 | 27,065 |
| No. of person-years | | 46,172 | 136,658 |
| *Total first primary cancers* | |  |  |
| No. of cases | | 387 | 830 |
| IR^a^ | | 838.2(754.7–921.7) | 607.4(566.0–648.7) |
| HR (95% CI)^b^ | Model 1^c^ | 1.29(1.13–1.48) | 1(ref) |
|  | Model 2^d^ | 1.27(1.10–1.46) | 1(ref) |
| *ORCs* | |  |  |
| No. of cases | | 147 | 330 |
| IR^a^ | | 318.4(266.9–369.8) | 241.5(215.4–267.5) |
| HR (95% CI)^b^ | Model 1^c^ | 1.42(1.13–1.79) | 1(ref) |
|  | Model 2^d^ | 1.39(1.10–1.76) | 1(ref) |
| *Total cancers minus ORCs* | |  |  |
| No. of cases | | 240 | 500 |
| IR^a^ | | 519.8(454.0–585.6) | 365.9(333.8–397.9) |
| HR (95% CI)^b^ | Model 1^c^ | 1.29(1.08–1.55) | 1(ref) |
|  | Model 2^d^ | 1.28(1.07–1.53) | 1(ref) |
| *Digestive tract organ cancers* | |  |  |
| No. of cases | | 86 | 212 |
| IR ^a^ | | 186.3(146.9–225.6) | 155.1(134.2–176.0) |
| HR (95% CI) ^b^ | Model 1^c^ | 1.07(0.79–1.44) | 1(ref) |
|  | Model 2^d^ | 1.03(0.76–1.39) | 1(ref) |
| *Digestive accessory organ cancers* | |  |  |
| No. of cases | | 50 | 71 |
| IR^a^ | | 108.3(78.3–138.3) | 52.0(39.9–64.0) |
| HR (95% CI)^b^ | Model 1^c^ | 1.92(1.27–2.88) | 1(ref) |
|  | Model 2^d^ | 2.05(1.34–3.12) | 1(ref) |
| *ORCs in reproductive organs* | |  |  |
| No. of cases | | 47 | 140 |
| IR^a^ | | 101.8(72.7–130.9) | 102.4(85.5–119.4) |
| HR (95% CI)^b^ | Model 1^c^ | 1.12(0.78–1.61) | 1(ref) |
|  | Model 2^d^ | 1.10(0.76–1.59) | 1(ref) |

Abbreviations: ORCs, obesity-related cancers; IR, incidence rate; HR, hazard ratio.

^a^ Per 100,000 person-years. ^b^ The HR for follow-up time window over 12 months was presented. ^c^ Model 1 was stratified on the matched set. ^d^ Model 2 was stratified on the matched set and further adjusted for waist circumference, physical activity, highest education level, annual household income, and cirrhosis/chronic hepatitis history (for composite cancer risks that encompassed liver cancer).

### Supplementary Table 13: Associations of new-onset T2DM with site-specific cancer risks in smoking- and alcohol-naïve women

|  | | **Smoking- and alcohol-naïve women** | |
| --- | --- | --- | --- |
|  |  | **New-onset T2DM** | **Matched unexposed** |
| No. of participants | | 9,052 | 27,065 |
| No. of person-years | | 46,172 | 136,658 |
| *Liver* | |  |  |
| No. of cases | | 30 | 52 |
| IR^a^ | | 65.0(41.7–88.2) | 38.1(27.7–48.4) |
| HR (95% CI)^b^ | Model 1^c^ | 1.68(1.02–2.78) | 1(ref) |
|  | Model 2^d^ | 1.83(1.08–3.12) | 1(ref) |
| *Colorectum* | |  |  |
| No. of cases | | 47 | 107 |
| IR^a^ | | 101.8(72.7–130.9) | 78.3(63.5–93.1) |
| HR (95% CI)^b^ | Model 1^c^ | 1.11(0.74–1.66) | 1(ref) |
|  | Model 2^d^ | 1.06(0.70–1.60) | 1(ref) |
| *Lung* | |  |  |
| No. of cases | | 61 | 116 |
| IR^a^ | | 132.1(99.0–165.3) | 84.9(69.4–100.3) |
| HR (95% CI)^b^ | Model 1^c^ | 1.30(0.91–1.87) | 1(ref) |
|  | Model 2^d^ | 1.26(0.87–1.82) | 1(ref) |
| *Stomach* | |  |  |
| No. of cases | | 30 | 67 |
| IR^a^ | | 65.0(41.7–88.2) | 49.0(37.3–60.8) |
| HR (95% CI)^b^ | Model 1^c^ | 1.13(0.67–1.91) | 1(ref) |
|  | Model 2^d^ | 1.07(0.63–1.82) | 1(ref) |
| *Breast (overall)* | |  |  |
| No. of cases | | 40 | 102 |
| IR^a^ | | 86.6(59.8–113.5) | 74.6(60.2–89.1) |
| HR (95% CI)^b^ | Model 1^c^ | 1.36(0.92–2.02) | 1(ref) |
|  | Model 2^d^ | 1.31(0.88–1.97) | 1(ref) |
| *Postmenopausal breast* | |  |  |
| No. of cases | | 37 | 97 |
| IR^a^ | | 80.1(54.3–106.0) | 71.0(56.9–85.1) |
| HR (95% CI)^b^ | Model 1^c^ | 1.31(0.87–1.98) | 1(ref) |
|  | Model 2^d^ | 1.28(0.84–1.94) | 1(ref) |

Abbreviations: IR, incidence rate; HR, hazard ratio.

^a^ Per 100,000 person-years. ^b^ The HR for follow-up time window over 12 months was presented. ^c^ Model 1 was stratified on the matched set. ^d^ Model 2 was stratified on the matched set and further adjusted for waist circumference, physical activity, highest education level, annual household income, and cirrhosis/chronic hepatitis history (for liver cancer).

### Supplementary Table 14: MR sensitivity analyses of the associations between T2DM and major site-specific cancers in East Asians

| **Cancer site & Source (No. of SNPs)** | **OR (95%CI)^a^** | **P** |
| --- | --- | --- |
| Colorectum & BBJ-CRC (183 SNPs) |  |  |
| IVW (P for heterogeneity=0.02) | 1.01 (0.97–1.05) | 0.62 |
| Weighted median | 0.97 (0.91–1.02) | 0.25 |
| Weighted mode | 0.95 (0.89–1.02) | 0.12 |
| MR-Egger (P for pleiotropy=0.13) | 0.96 (0.89–1.03) | 0.28 |
| MR-PRESSO (no outliers detected) | 1.01 (0.97–1.05) | 0.62 |
| Colorectum & CKB (185 SNPs) |  |  |
| IVW (P for heterogeneity=0.20) | 0.97(0.90–1.05) | 0.50 |
| Weighted median | 0.98(0.85–1.13) | 0.77 |
| Weighted mode | 0.96(0.80–1.15) | 0.67 |
| MR-Egger (P for pleiotropy=0.80) | 0.95(0.81–1.12) | 0.58 |
| MR-PRESSO (no outliers detected) | 0.97(0.90–1.05) | 0.50 |
| Pancreas & JaPAN+NCC+BBJ-PaC (170 SNPs) |  |  |
| IVW (P for heterogeneity=0.64) | 1.10(1.03–1.17) | 0.0033 |
| Weighted median | 1.13(1.02–1.26) | 0.03 |
| Weighted mode | 1.16(1.04–1.30) | 0.02 |
| MR-Egger (P for pleiotropy=0.47) | 1.14(1.01–1.30) | 0.03 |
| MR-PRESSO (no outliers detected) | 1.10(1.03–1.17) | 0.0033 |
| Pancreas & CKB (185 SNPs) |  |  |
| IVW (P for heterogeneity=0.65) | 0.97(0.82–1.14) | 0.67 |
| Weighted median | 0.90(0.68–1.18) | 0.44 |
| Weighted mode | 0.81(0.57–1.15) | 0.24 |
| MR-Egger (P for pleiotropy=0.14) | 0.78(0.57–1.08) | 0.14 |
| MR-PRESSO (no outliers detected) | 0.97(0.82–1.14) | 0.67 |
| Breast & KoGES (182 SNPs) |  |  |
| IVW (P for heterogeneity=0.04) | 1.05(0.93–1.19) | 0.40 |
| Weighted median | 1.09(0.91–1.32) | 0.35 |
| Weighted mode | 1.09(0.87–1.37) | 0.44 |
| MR-Egger (P for pleiotropy=0.02) | 0.82(0.64–1.04) | 0.10 |
| MR-PRESSO (no outliers detected) | 1.05(0.93–1.19) | 0.40 |
| Breast & CKB (185 SNPs) |  |  |
| IVW (P for heterogeneity= 0.79) | 1.04(0.95–1.14) | 0.37 |
| Weighted median | 1.08(0.93–1.25) | 0.36 |
| Weighted mode | 1.10(0.90–1.34) | 0.35 |
| MR-Egger (P for pleiotropy=0.50) | 0.99(0.82–1.19) | 0.89 |
| MR-PRESSO (no outliers detected) | 1.04(0.95–1.14) | 0.37 |
| Endometrium & KoGES (182 SNPs) |  |  |
| IVW (P for heterogeneity=0.31) | 0.97(0.86–1.10) | 0.62 |
| Weighted median | 1.08(0.87–1.34) | 0.47 |
| Weighted mode | 1.07(0.84–1.36) | 0.61 |
| MR-Egger (P for pleiotropy=0.42) | 1.06(0.83–1.36) | 0.65 |
| MR-PRESSO (no outliers detected) | 0.97(0.86–1.10) | 0.62 |
| Endometrium & CKB (185 SNPs) |  |  |
| IVW (P for heterogeneity=0.96) | 1.15(0.91–1.45) | 0.25 |
| Weighted median | 1.33(0.90–1.98) | 0.15 |
| Weighted mode | .. | .. |
| MR-Egger (P for pleiotropy=0.50) | 1.32(0.83–2.11) | 0.25 |
| MR-PRESSO (no outliers detected) | 1.15(0.91–1.45) | 0.25 |
| Liver & CKB (185 SNPs) |  |  |
| IVW (P for heterogeneity=0.06) | 1.00(0.91–1.09) | 0.97 |
| Weighted median | 1.10(0.94–1.28) | 0.21 |
| Weighted mode | 1.13(0.94–1.37) | 0.23 |
| MR-Egger (P for pleiotropy=0.44) | 1.06(0.88–1.28) | 0.52 |
| MR-PRESSO (no outliers detected) | 1.00(0.91–1.09) | 0.97 |
| Lung & CKB (185 SNPs) |  |  |
| IVW (P for heterogeneity=0.04) | 1.02(0.96–1.08) | 0.63 |
| Weighted median | 1.05(0.96–1.15) | 0.32 |
| Weighted mode | 1.06(0.95–1.17) | 0.32 |
| MR-Egger (P for pleiotropy=0.85) | 1.03(0.91–1.16) | 0.69 |
| MR-PRESSO (no outliers detected) | 1.02(0.96–1.08) | 0.63 |
| Stomach & CKB (185 SNPs) |  |  |
| IVW (P for heterogeneity=0.76) | 1.00(0.92–1.08) | 0.99 |
| Weighted median | 0.90(0.79–1.03) | 0.13 |
| Weighted mode | 0.91(0.79–1.06) | 0.23 |
| MR-Egger (P for pleiotropy=0.93) | 0.99(0.85–1.17) | 0.94 |
| MR-PRESSO (no outliers detected) | 1.00(0.92–1.08) | 0.99 |

Abbreviations: T2DM, type 2 diabetes mellitus; BBJ-CRC, Biobank Japan colorectal cancer cohort; CKB, China Kadoorie Biobank; JaPAN, Japan pancreatic cancer research; NCC, national cancer centre; BBJ-PaC, Biobank Japan pancreatic cancer cohort; KoGES, Korean genome and epidemiology study; OR, odds ratio; SNPs, single nucleotide polymorphisms.

^a^ Estimates indicating OR of cancer per doubling in odds of T2DM.

### Supplementary Table 15: Power calculation for two-sample Mendelian randomisation of the associations between T2DM and site-specific cancers^a^

| **Cancer site& source** | **Sample size** | **No. of case** | **No. of control** | **Case-control ratio** | **Odds ratio^b^** | | |
| --- | --- | --- | --- | --- | --- | --- | --- |
|  |  |  |  |  | **0.90** | **1.10** | **1.20** |
| Colorectum |  |  |  |  |  |  |  |
| BBJ-CRC | 33,870 | 6,692 | 27,178 | 4.06 | 1.00 | 0.99 | 1.00 |
| CKB | 100,640 | 912 | 99,728 | 109.35 | 0.51 | 0.43 | 0.93 |
| Pancreas |  |  |  |  |  |  |  |
| JaPAN+NCC+BBJ-PaC | 34,631 | 2,039 | 32,592 | 15.98 | 0.82 | 0.74 | 1.00 |
| CKB | 100,640 | 205 | 100,435 | 489.93 | 0.15 | 0.13 | 0.37 |
| Breast |  |  |  |  |  |  |  |
| KoGES | 46,330 | 424 | 45,906 | 108.27 | 0.27 | 0.23 | 0.65 |
| CKB | 57,625 | 655 | 56,970 | 86.98 | 0.39 | 0.33 | 0.83 |
| Endometrium |  |  |  |  |  |  |  |
| KoGES | 46,330 | 355 | 45,975 | 129.51 | 0.23 | 0.20 | 0.57 |
| CKB | 57,625 | 102 | 57,523 | 563.95 | 0.10 | 0.09 | 0.21 |
| Liver |  |  |  |  |  |  |  |
| CKB | 100,640 | 770 | 99,870 | 129.70 | 0.44 | 0.38 | 0.88 |
| Lung |  |  |  |  |  |  |  |
| CKB | 100,640 | 1,802 | 98,838 | 54.85 | 0.79 | 0.71 | 1.00 |
| Stomach |  |  |  |  |  |  |  |
| CKB | 100,640 | 888 | 99,752 | 112.33 | 0.50 | 0.42 | 0.92 |

Abbreviations: T2DM, type 2 diabetes mellitus; BBJ-CRC, Biobank Japan colorectal cancer cohort; CKB, China Kadoorie Biobank; JaPAN, Japan pancreatic cancer research; NCC, national cancer centre; BBJ-PaC, Biobank Japan pancreatic cancer cohort; KoGES, Korean genome and epidemiology study; OR, odds ratio.

^a^ R^2^ equal to 0.389 (corresponding to 193 SNPs) was applied for calculations in this table. ^b^ OR of cancer per one unit increase in log-odds of T2DM.

### Supplementary Table 16: Triangulation of evidence from matched-cohort analysis in CKB and MR analysis in East Asians for the casual effect of T2DM on site-specific cancer risks

| **Cancer site** | **Effect estimates** | | | **Interpretation** | **Bias diagnoses** |
| --- | --- | --- | --- | --- | --- |
|  | OR^a^ | HR^b^ | |  |  |
|  |  | Men | Women |  |  |
| *Pancreas* | 1.08(1.02–1.15 | 2.57(1.35–4.92) | 3.95(1.92–8.13) | Concordant and precise | Residual confounding from alcohol related exposures could result in inflated effect magnitude in HR. |
| *Liver* | 1.00(0.91–1.09) | 2.12(1.42–3.15) | 2.39(1.42–4.04) | Discordant | Restricted power in MR could lead to the failure to detect a small effect; residual confounding from major contributors to incident liver cancer in China (e.g., HBV/HCV infections) could result in inflated effect magnitude in HR. |
| *Colorectum* | 1.00(0.97–1.04) | 1.33(0.93–1.91) | 1.16(0.82–1.64) | Concordant and precise | Residual confounding from alcohol related exposures could result in inflated effect magnitude in HR. |
| *Breast* | 1.05(0.97–1.13) | - | 1.01(0.73–1.38)^c^ | Concordant but imprecise | Limited cancer case number in MR analyses increases imprecision in effect estimates, reflected by a wide 95% CI across 1. |
| *Lung* | 1.02(0.96–1.08) | 1.55(1.17–2.05) | 1.15(0.87–1.52) | Discordant | The only inconsistency came from a meaningful HR estimate across 1 in men, which could be biased from smoking-related confounding; sex-specific MR and (or) factorial MR testing for exposure–smoking interactions when power allows could give new insights into this issue. |
| *Stomach* | 1.00(0.92–1.08) | 1.13(0.77–1.68) | 1.11(0.72–1.71) | Concordant but imprecise | Limited cancer case number in both MR and observational analyses increases imprecision in effect estimates, reflected by a wide 95% CI across 1. |

Abbreviations: OR, odds ratio; HR, hazard ratio; HBV, hepatitis B virus; HCV, hepatitis C virus.

^a^ MR estimate indicating the OR in cancer per doubling in odds of T2DM; ^b^ Observational estimate from the multivariable-adjusted Cox model following 1:3 matching, indicating the HR in cancer in new-onset T2DM vs matched control. ^c^ HR for overall breast cancer in women.

Interpretation categories: (i) Concordant and precise: same effect direction with precise estimates in both methods.; (ii) Concordant but imprecise: same effect direction with imprecise estimates in either method; (iii) Discordant: different effect directions in the two methods.

### Supplementary Figure 1: Flowchart showing the confirmation of prevalent diabetes cases

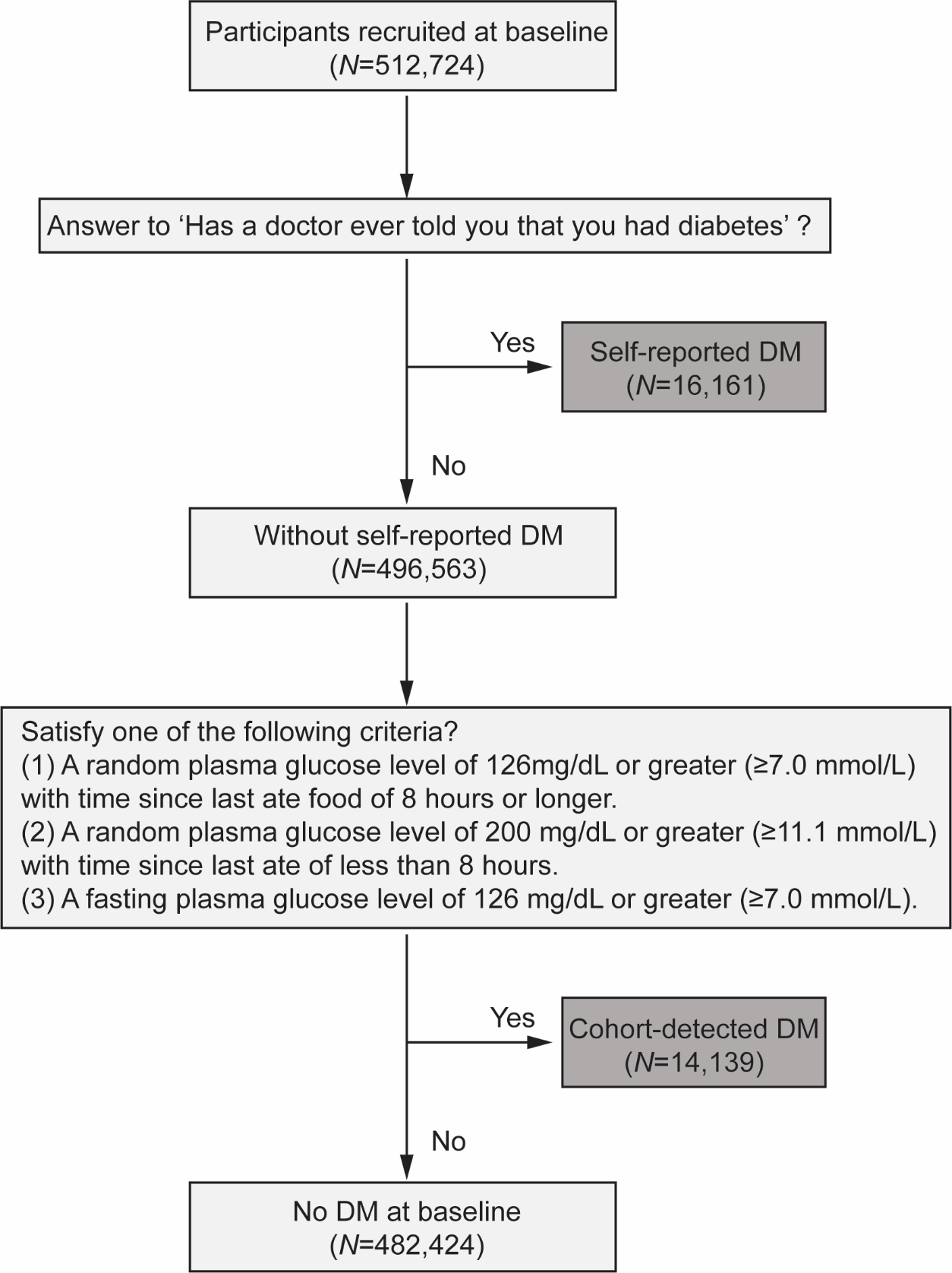

Abbreviations: DM, diabetes mellitus.

Those participants who gave a ‘YES’ response to the question ‘Has a doctor EVER told you that you had diabetes?’ were counted into ‘self-reported diabetes’. At the same time, those gave a ‘NO’ answer to the previous question, but satisfy one of the following criteria were regarded as ‘cohort-detected diabetes’: (1) a random plasma glucose level of 126 mg/dL or greater (≥7.0 mmol/L) with time since last ate food of 8 hours or longer; (2) a random plasma glucose level of 200 mg/dL or greater (≥11.1 mmol/L) with time since last ate of less than 8 hours; (3) a fasting plasma glucose level of 126 mg/dL or greater (≥7.0 mmol/L).

### Supplementary Figure 2: Illustration of the longitudinal matching design in the present study

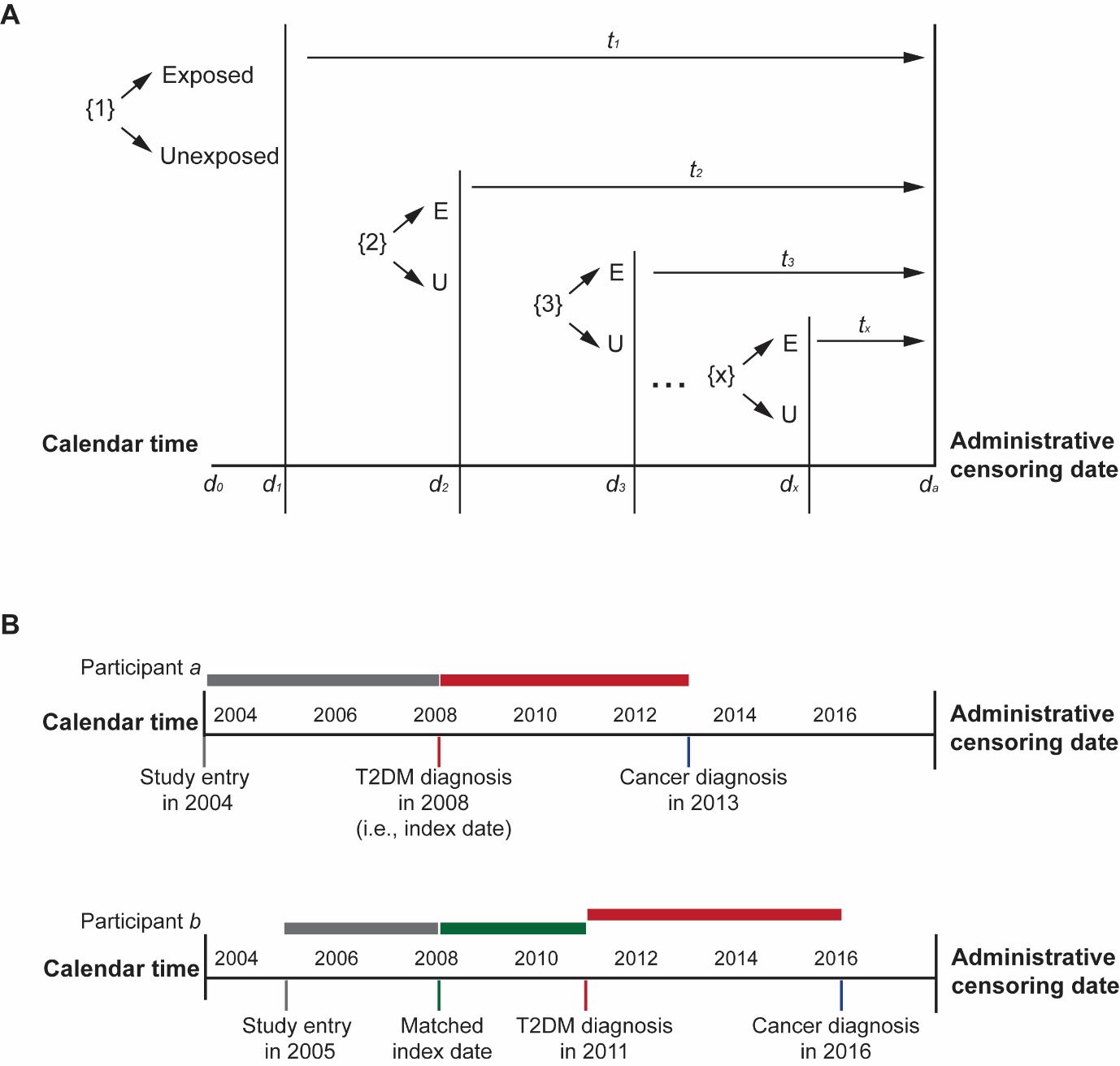

**(A)** The schema of sequential allocations of participants to the exposed or unexposed group. The relevant time scale for matching was the calendar time (*d*). Follow-up for outcomes (time scale *t*) began after the assigned index date *d_x_*. The possible maximum follow-up time for every participant was the time lag between *d_a_* and *d_x_*, where *d_a_* was the administrative censoring date. **(B)** Illustration of two hypothetical participants in a matched pair, where participant ***a*** was the initially exposed individual and participant ***b*** the matched unexposed. A *per-protocol* analysis is presented here**,** where participant ***b*** was followed up as unexposed (indicated by green bar) from the assigned index date to a new diagnosis of T2DM in this matched pair, and in another stratum, as an exposed individual with new follow-up since T2DM diagnosis (indicated by red bar).

### Supplementary Figure 3: Illustration of the study design in Mendelian randomisation

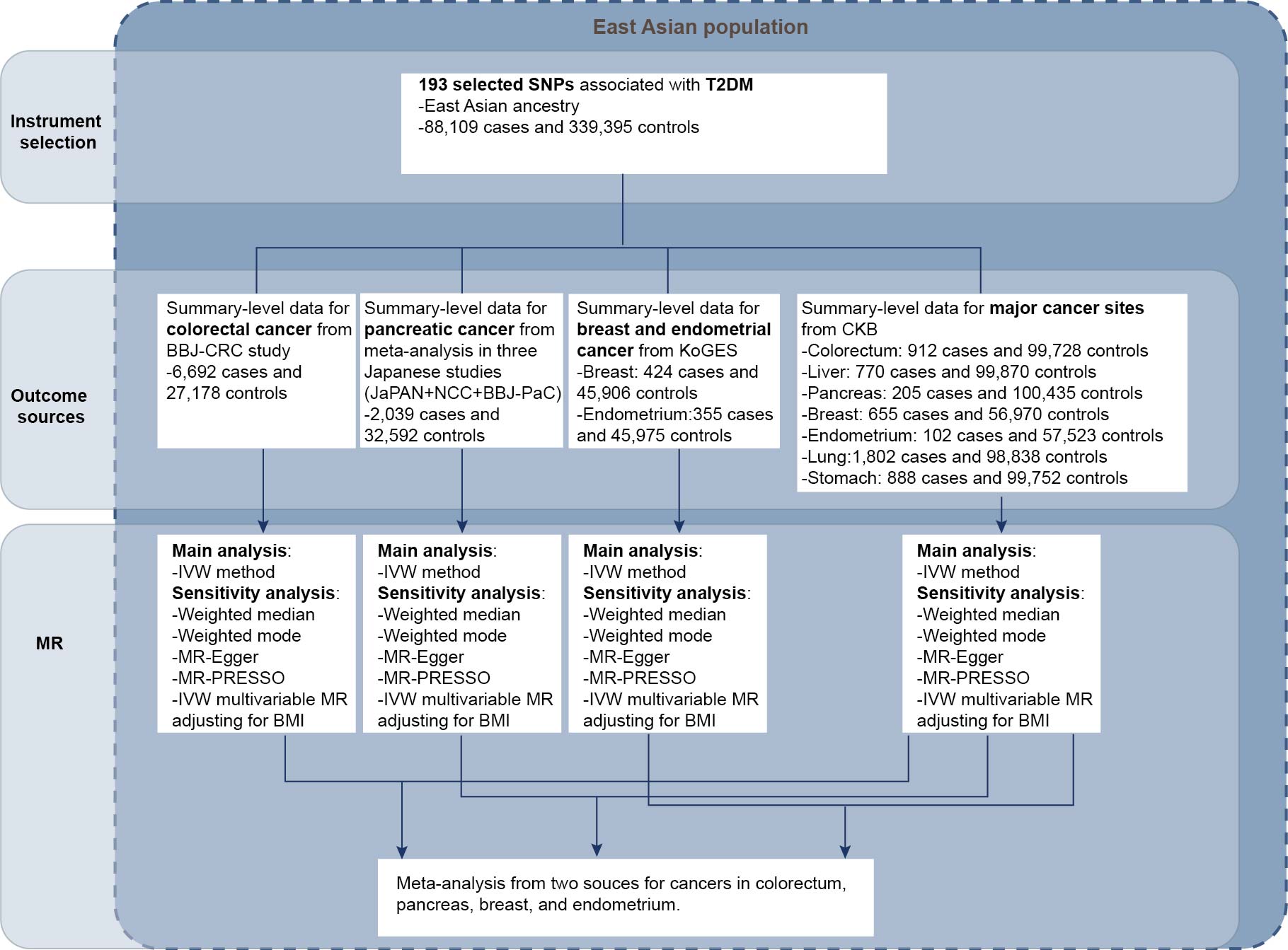

Abbreviations: SNPs, single nucleotide polymorphisms; MR, Mendelian randomisation; T2DM, type 2 diabetes mellitus; BBJ-CRC, Biobank Japan colorectal cancer cohort; CKB, China Kadoorie Biobank; JaPAN, Japan pancreatic cancer research; NCC, National Cancer Centre; BBJ-PaC, Biobank Japan pancreatic cancer cohort; KoGES, Korean Genome and Epidemiology Study; IVW, inverse-variance weighted; MR-PRESSO, MR Pleiotropy RESidual Sum and Outlier methods.

### Supplementary Figure 4: Flowchart for identifying eligible individuals for matching at baseline

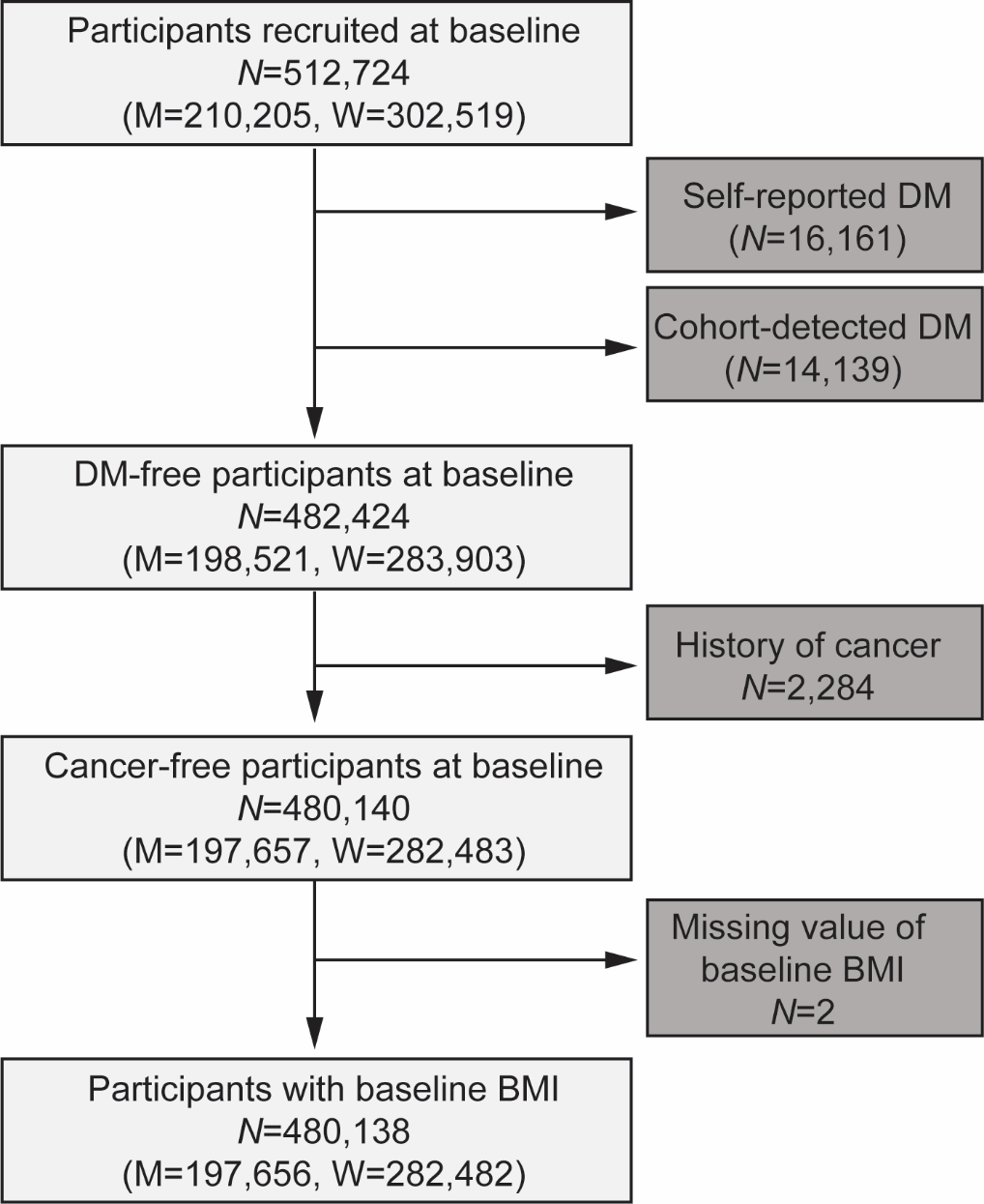

Abbreviations: DM, diabetes mellitus; BMI, body mass index; M, men; W, women.

### Supplementary Figure 5: Age-standardised incidence rate of T2DM in men and women across the ten study regions

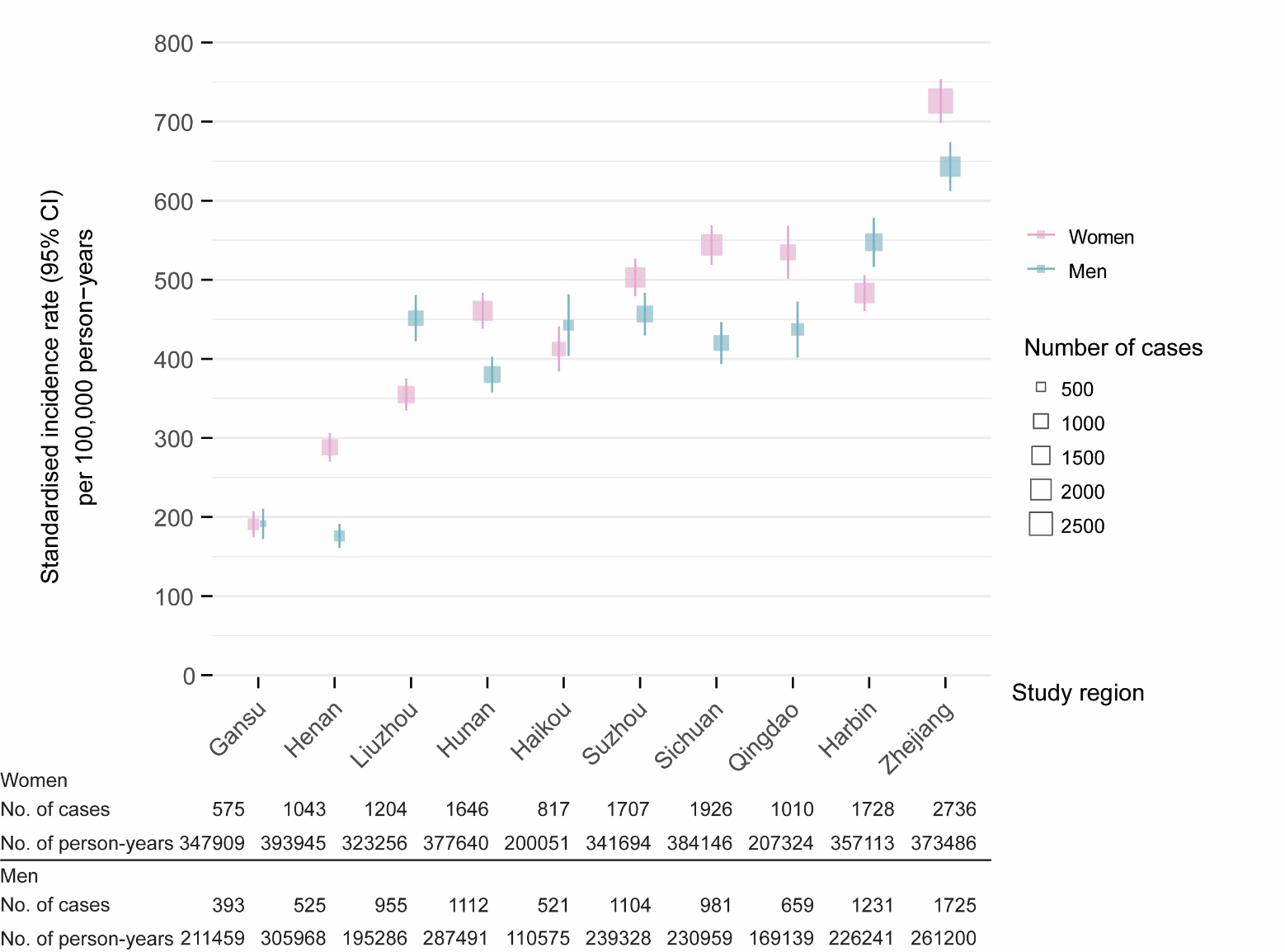

T2DM risks (per 100,000 person-years) were standardised by baseline age (<40/40–49/50–59/60–69/≥70 years) using the total diabetes-free population at baseline in CKB as the standard. The size of each box is proportional to the number of new-onset T2DM cases in each group and error bars indicate the 95% confidence interval estimated by assuming a Poisson distribution.

### Supplementary Figure 6: Distribution of age at T2DM diagnosis among participants eligible for matching as exposed individuals

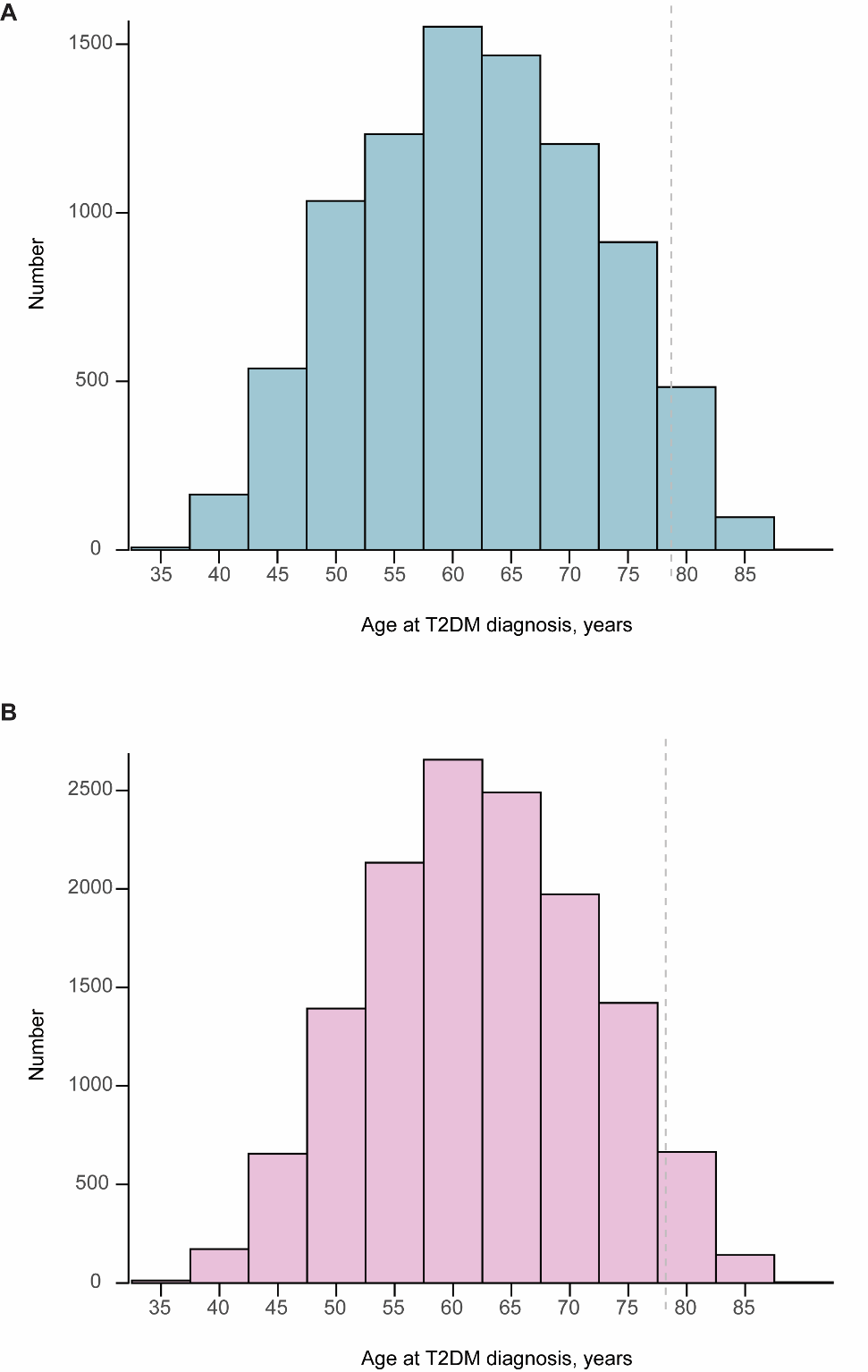

**(A)** Men. **(B)** Women. The 95^th^ percentiles, indicated by grey dashed lines, are 78.7 years for men and 78.2 years for women.

### Supplementary Figure 7: Distribution of baseline body mass index among participants eligible for matching as exposed individuals

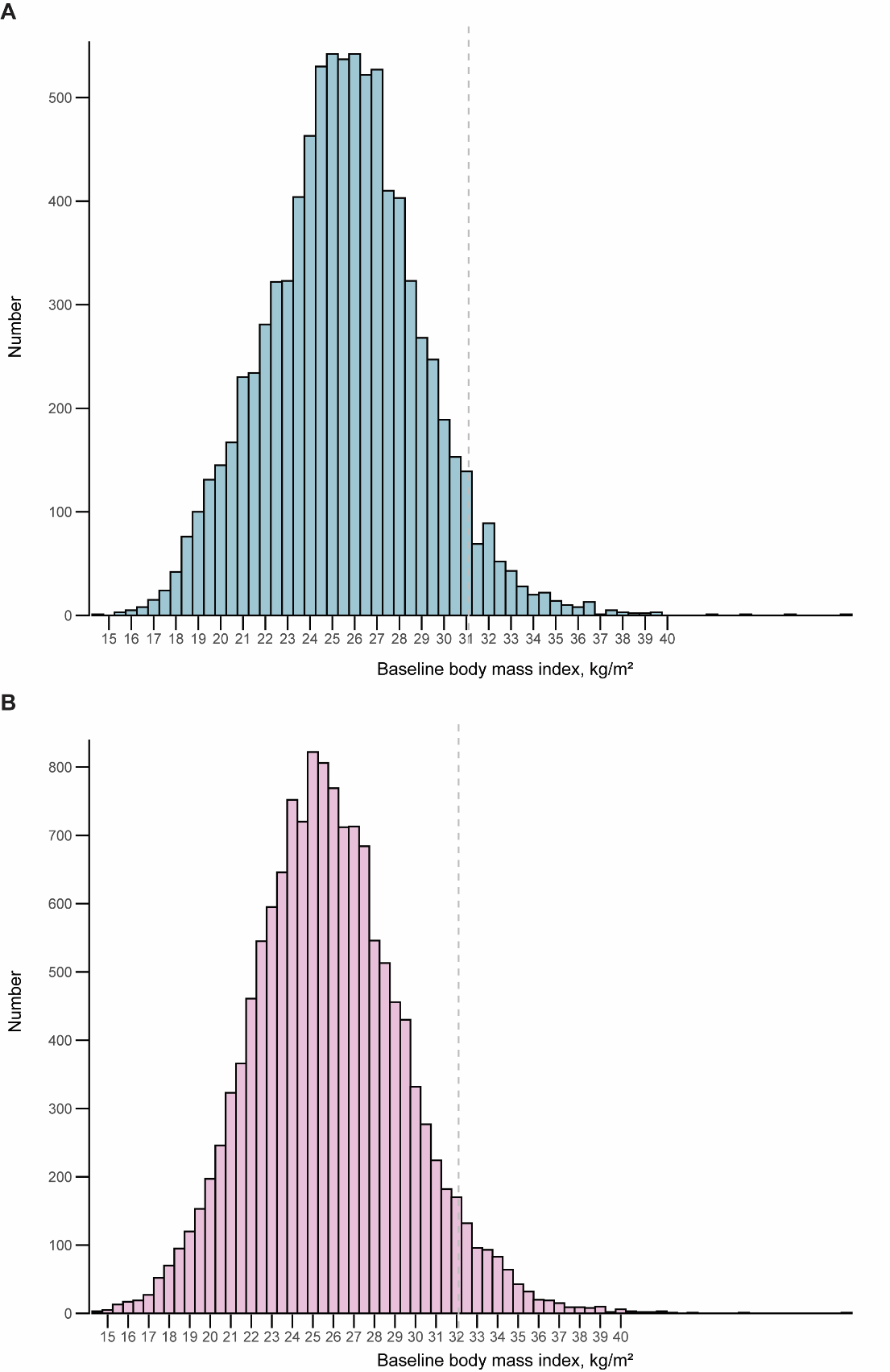

**(A)** Men. **(B)** Women. The 95^th^ percentiles, indicated by grey dashed lines, are 31.1 kg/m^2^ for men and 32.1 kg/m^2^ for women.

### Supplementary Figure 8: Schoenfeld residuals for T2DM status from a standard Cox model with constant coefficients

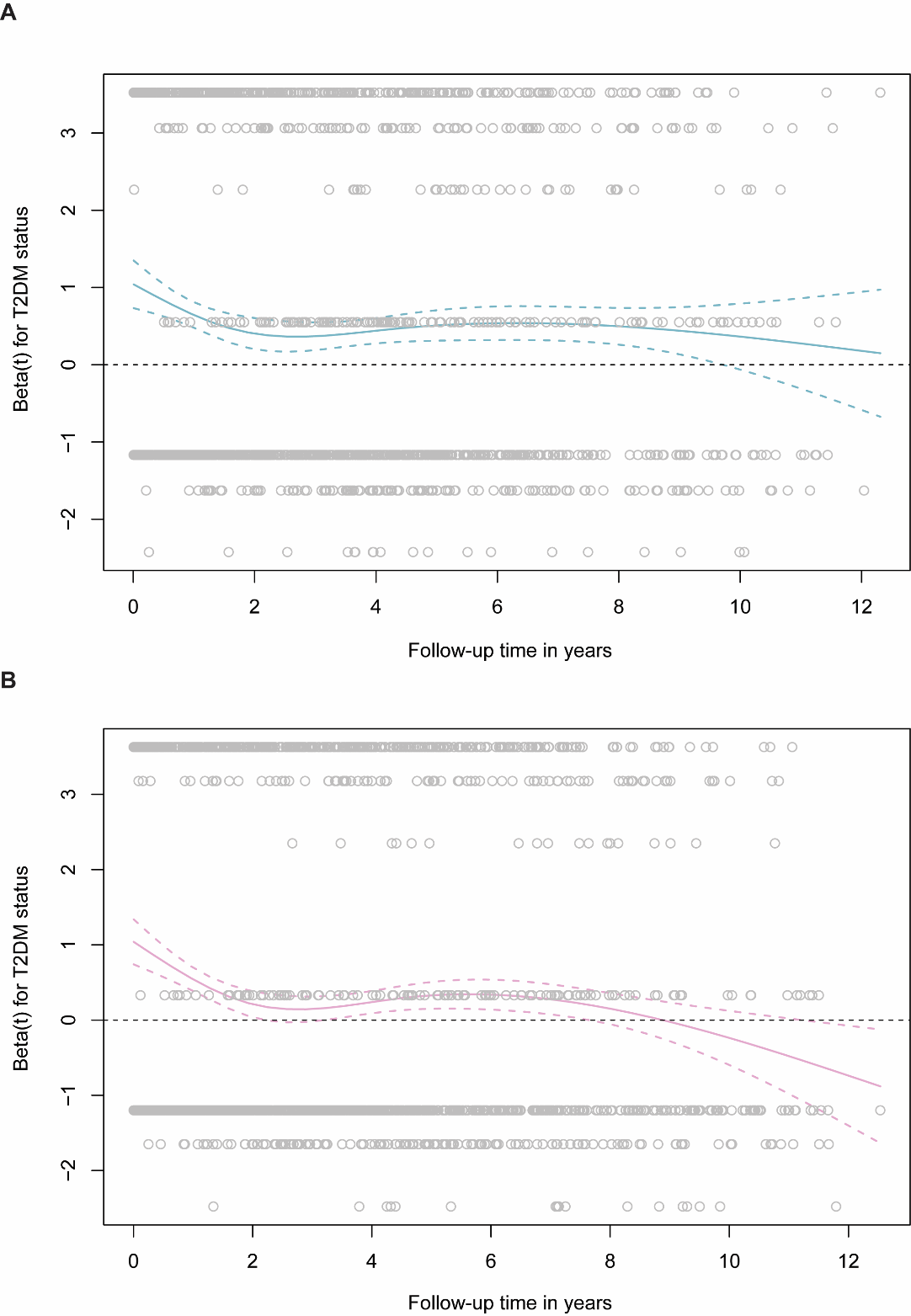

**(A)** Men. **(B)** Women. Residuals are plotted against follow-up time (years). Solid lines represent the fitted trend, and dashed lines denote the 95% confidence interval.

### Supplementary Figure 9: Cumulative incidence function for total first primary cancers and competing non-cancer deaths

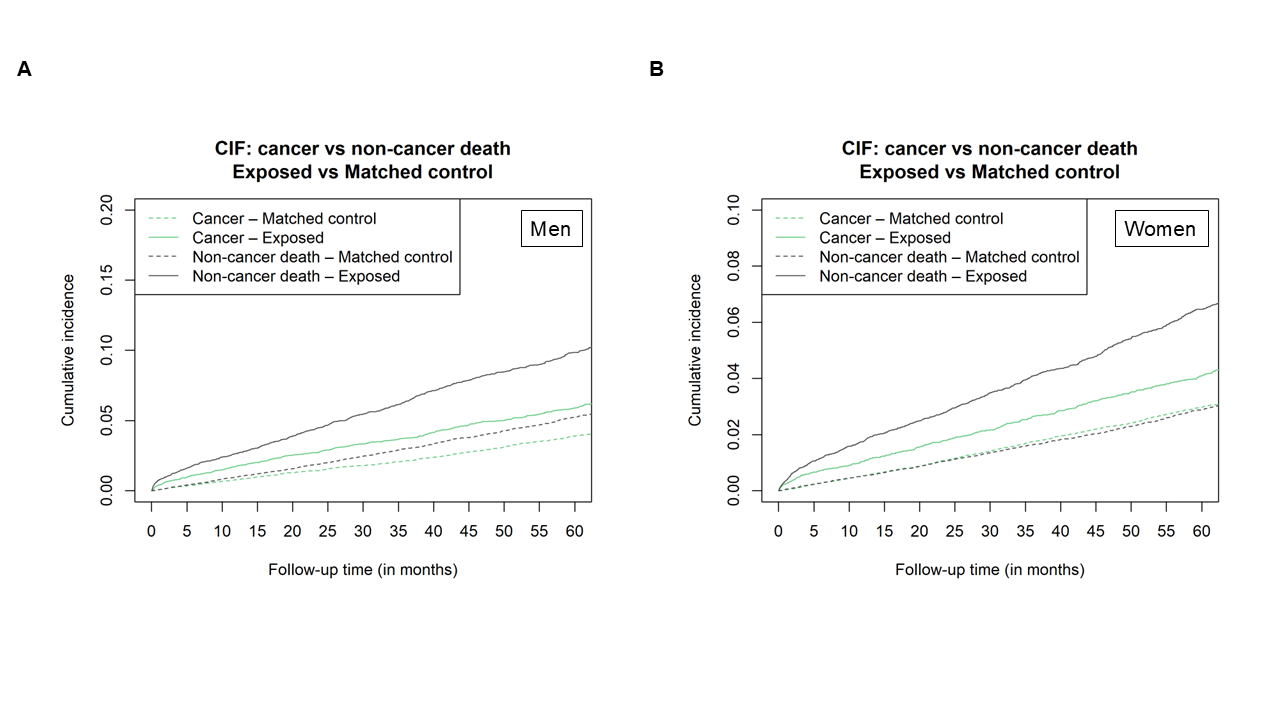

**(A)** Men. **(B)** Women.

The y-axis indicates the probability, and x-axis only shows the follow-up time window within 60-month

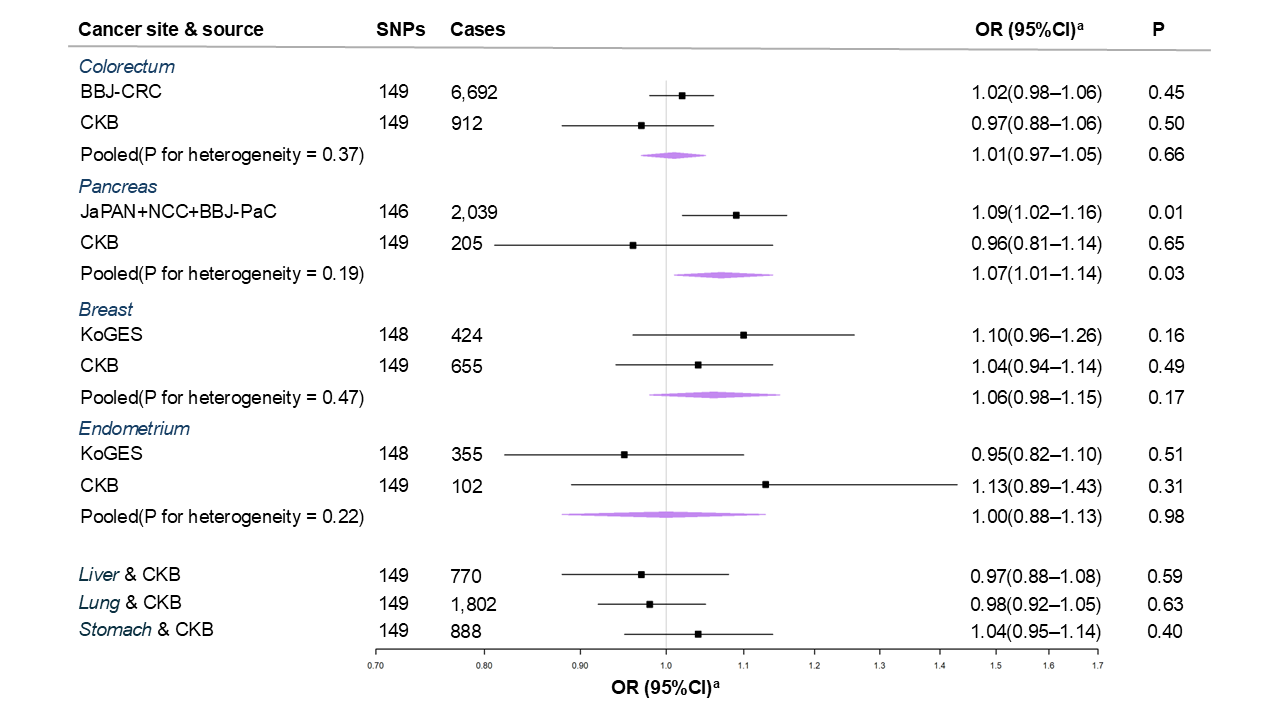

### Supplementary Figure 10: Multivariable MR with adjustments for BMI of the associations of T2DM with major site-specific cancers in East Asians

Abbreviations: T2DM, type 2 diabetes mellitus; BMI, body mass index; BBJ-CRC, Biobank Japan colorectal cancer cohort; CKB, China Kadoorie Biobank; JaPAN, Japan pancreatic cancer research; NCC, national cancer centre; BBJ-PaC, Biobank Japan pancreatic cancer cohort; KoGES, Korean genome and epidemiology study; OR, odds ratio; SNPs, single nucleotide polymorphisms.

^a^ Estimates indicating OR of cancer per doubling in odds of T2DM.
